# Heterogeneity in transmissibility and shedding SARS-CoV-2 via droplets and aerosols

**DOI:** 10.1101/2020.10.13.20212233

**Authors:** Paul Z. Chen, Niklas Bobrovitz, Zahra Premji, Marion Koopmans, David N. Fisman, Frank X. Gu

**Author notes:** To whom correspondence may be addressed: Frank X. Gu. **Email:**.

## Abstract

Which virological factors mediate overdispersion in the transmissibility of emerging viruses remains a longstanding question in infectious disease epidemiology. Here, we use systematic review to develop a comprehensive dataset of respiratory viral loads (rVLs) of SARS-CoV-2, SARS-CoV-1 and influenza A(H1N1)pdm09. We then comparatively meta-analyze the data and model individual infectiousness by shedding viable virus via respiratory droplets and aerosols. Our analyses indicate heterogeneity in rVL as an intrinsic virological factor facilitating greater overdispersion for SARS-CoV-2 in the COVID-19 pandemic than A(H1N1)pdm09 in the 2009 influenza pandemic. For COVID-19, case heterogeneity remains broad throughout the infectious period, including for pediatric and asymptomatic infections. Hence, many COVID-19 cases inherently present minimal transmission risk, whereas highly infectious individuals shed tens to thousands of SARS-CoV-2 virions/min via droplets and aerosols while breathing, talking and singing. Coughing increases the contagiousness, especially in close contact, of symptomatic cases relative to asymptomatic ones. Infectiousness tends to be elevated between 1-5 days post-symptom onset. Our findings show how individual case variations influence virus transmissibility and present considerations for disease control in the COVID-19 pandemic.

**Significance Statement:** For some emerging infectious diseases, including COVID-19, few cases cause most secondary infections. Others, like influenza A(H1N1)pdm09, spread more homogenously. The virological factors that mediate such distinctions in transmissibility remain unelucidated, prohibiting the development of specific disease control measures. We find that intrinsic case variation in respiratory viral load (rVL) facilitates overdispersion, and superspreading, for COVID-19 but more homogeneous transmission for A(H1N1)pdm09. We interpret the influence of heterogeneity in rVL on individual infectiousness by modelling likelihoods of shedding viable virus via respiratory droplets and aerosols. We analyze the distribution and kinetics of SARS-CoV-2 rVL, including across age and symptomatology subgroups. Our findings compare individual infectiousness across COVID-19 and A(H1N1)pdm09 cases and present quantitative guidance on triaging COVID-19 contact tracing.

## Main Text

Severe acute respiratory syndrome coronavirus 2 (SARS-CoV-2) has spread globally, causing the coronavirus disease 2019 (COVID-19) pandemic with more than 62.5 million infections and 1.4 million deaths (as of 29 November 2020) (1). For respiratory virus transmission, airway epithelial cells shed virions to the extracellular fluid before atomization (from breathing, talking, singing, coughing and aerosol-generating procedures) partitions them into a polydisperse mixture of particles that are expelled to the ambient environment. Aerosols (≤100 μm) can be inhaled nasally, whereas droplets (>100 μm) tend to be excluded (2, 3). For direct transmission, droplets must be sprayed ballistically onto susceptible tissue (4). Hence, droplets predominantly deposit on nearby surfaces, potentiating indirect transmission. Aerosols can be further categorized based on typical travel characteristics: short-range aerosols (50-100 μm) tend to settle within 2 m; long-range ones (10-50 μm) often travel beyond 2 m based on emission force; and buoyant aerosols (≤10 μm) remain suspended and travel based on airflow profiles for minutes to many hours (4, 5). Although proximity has been associated with infection risk for COVID-19 (6), studies have also suggested that long-range airborne transmission occurs conditionally (7-9).

While the basic reproductive number has been estimated to be 2.0-3.6 (10, 11), transmissibility of SARS-CoV-2 is highly overdispersed (dispersion parameter *k*, 0.10-0.58), with numerous instances of superspreading (7-9) and few cases (10-20%) causing many secondary infections (80%) (12-14). Similarly, few cases drive the transmission of SARS-CoV-1 (*k*, 0.16-0.17) (15), whereas influenza A(H1N1)pdm09 transmits more homogeneously (*k*, 7.4-14.4) (16, 17), despite both viruses spreading by contact, droplets and aerosols (18, 19). Although understanding the determinants of viral overdispersion is crucial towards characterizing transmissibility and developing effective strategies to limit infection (20), mechanistic associations for *k* remain unclear. As an empirical estimate, *k* depends on myriad extrinsic (behavioral, environmental and invention) and host factors. Nonetheless, *k* remains similar across distinct outbreaks (15), suggesting that intrinsic virological factors mediate overdispersion for emerging viruses.

Here, we investigated how intrinsic case variation in rVL facilitates overdispersion in SARS-CoV-2 transmissibility. By systematic review, we developed a comprehensive dataset of rVLs from cases of COVID-19, SARS and A(H1N1)pdm09. Using comparative meta-analyses, we found that heterogeneity in rVL was associated with overdispersion among these emerging infections. To assess potential sources of case heterogeneity, we analyzed SARS-CoV-2 rVLs across age and symptomatology subgroups as well as disease course. To interpret the influence of heterogeneity in rVL on individual infectiousness, we modelled likelihoods of shedding viable virus via respiratory droplets and aerosols.

## Results

### Systematic review

We conducted a systematic review on virus quantitation in respiratory specimens taken during the infectious periods of SARS-CoV-2 (−3 to 10 days from symptom onset [DFSO]) (21-23), SARS-CoV-1 (0-20 DFSO) (24) and A(H1N1)pdm09 (−2 to 9 DFSO) (25) (Methods). The systematic search (*SI Appendix*, Tables S1 to S5) identified 4,274 results. After screening and full-text review, 64 studies met the inclusion criteria and contributed to the systematic dataset (Fig. 1) (*N* = 9,631 total specimens), which included adult (*N* = 5,033) and pediatric (*N* = 1,608) cases from 15 countries and specimen measurements for asymptomatic (*N* = 2,387), presymptomatic (*N* = 28) and symptomatic (*N* = 7,161) infections. According to a hybrid Joanna Briggs Institute critical appraisal checklist, risk of bias was low for most contributing studies (*SI Appendix*, Table S6).

**Fig. 1.**
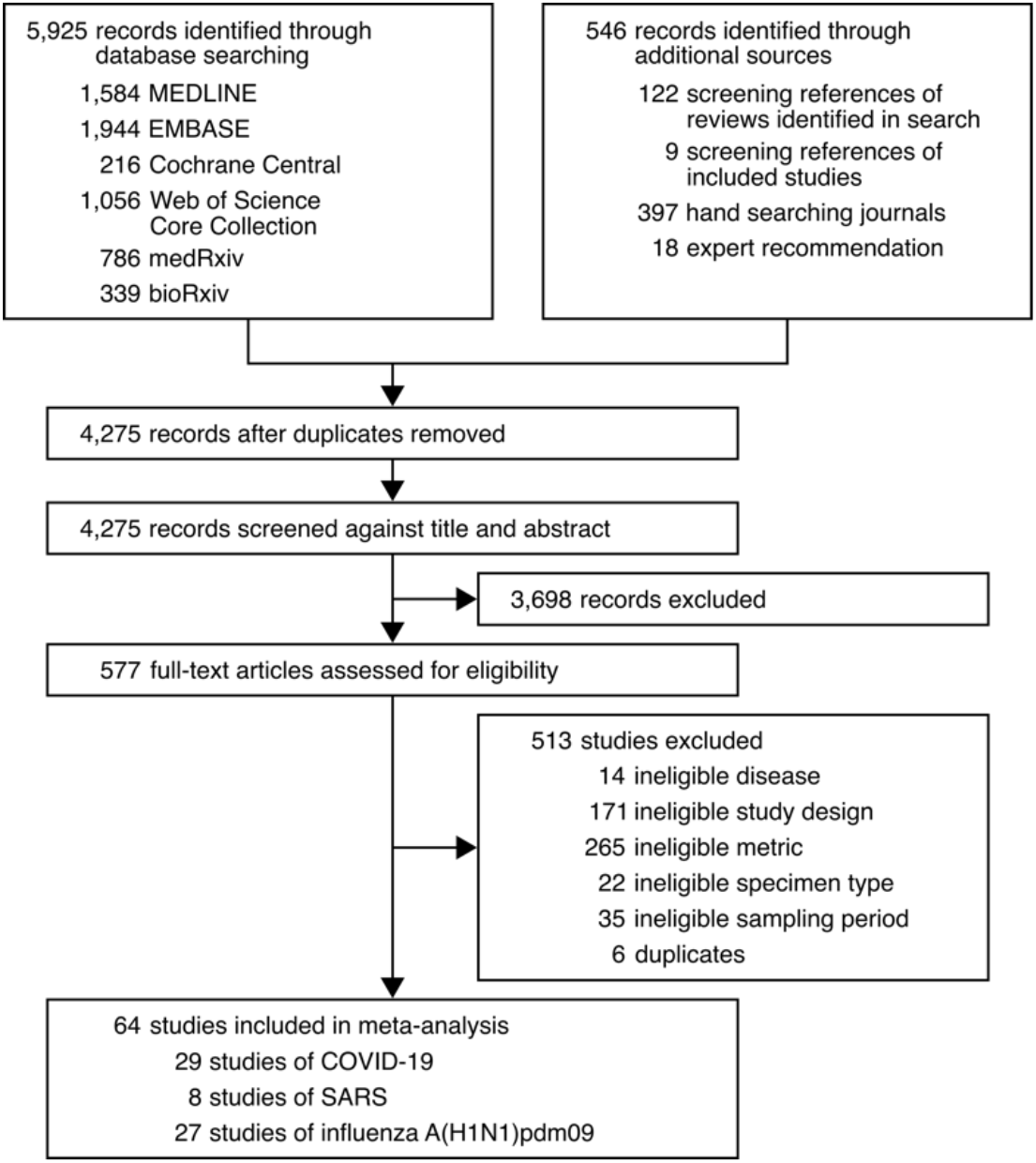
Development of the systematic dataset.

### Association of overdispersion with heterogeneity in rVL

We hypothesized that individual case variation in rVL facilitates the distinctions in *k* among COVID-19, SARS and A(H1N1)pdm09. For each study in the systematic dataset, we used specimen measurements (viral RNA concentration in a respiratory specimen) to estimate rVLs (viral RNA concentration in the respiratory tract) (Methods). To investigate the relationship between *k* and heterogeneity in rVL, we performed a meta-regression using each contributing study (*SI Appendix*, Fig. S1), which showed a weak, negative association between the two variables (meta-regression slope *t*-test: *P* = 0.038, Pearson’s *r* = -0.26).

Using contributing studies with low risk of bias, meta-regression (Fig. 2) showed a strong, negative association between *k* and heterogeneity in rVL (meta-regression slope *t*-test: *P* < 10^−9^, Pearson’s *r* = -0.73). In this case, each unit increase (1 log_10_ copies/ml) in the standard deviation (SD) of rVL decreased log(*k*) by a factor of -1.41 (95% confidence interval [CI]: -1.78 to -1.03), suggesting that broader heterogeneity in rVL facilitates greater overdispersion in the transmissibility of SARS-CoV-2 than of A(H1N1)pmd09. To investigate mechanistic aspects of this association, we conducted a series of analyses on rVL and then modelled the influence of heterogeneity in rVL on individual infectiousness.

**Fig. 2.**
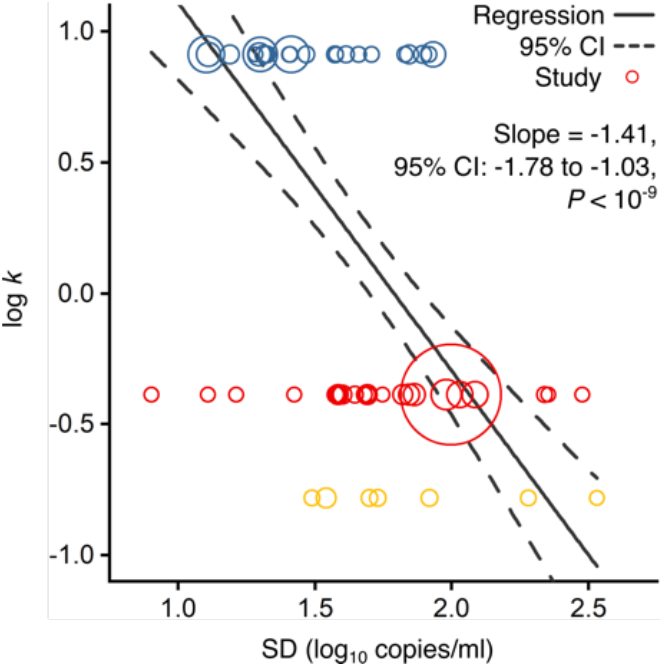
Association of overdispersion in SARS-CoV-2, SARS-CoV-1 and A(H1N1)pdm09 transmissibility with heterogeneity in rVL. Meta-regression of dispersion parameter (*k*) with the standard deviation (SD) of respiratory viral loads (rVLs) from contributing studies with low risk of bias (Pearson’s *r* = -0.73). Pooled estimates of k were determined from the literature for each infection. Blue, red and yellow circles denote A(H1N1)pdm09 (*N* = 22), COVID-19 (*N* = 24) and SARS (*N* = 7) studies, respectively. Circle sizes denote weighting in the meta-regression. The *P*-value was obtained using the meta-regression slope *t*-test.

### Meta-analysis and subgroup analyses of rVL

We first compared rVLs among the emerging infections. We performed a random-effects meta-analysis (*SI Appendix*, Fig. S2), which assessed the expected rVL when encountering a COVID-19, SARS or A(H1N1)pdm09 case during the infectious period. This showed that the expected rVL of SARS-CoV-2 was comparable to that of SARS-CoV-1 (one-sided Welch’s *t*-test: *P* = 0.111) but lesser than that of A(H1N1)pdm09 (*P* = 0.040). We also performed random-effects subgroup analyses for COVID-19 (Fig. 3), which showed that expected SARS-CoV-2 rVLs were similar between pediatric and adult cases (*P* = 0.476) and comparable between symptomatic/presymptomatic and asymptomatic infections (*P* = 0.090). Since these meta-analyses had significant between-study heterogeneity among the mean estimates (Cochran’s *Q* test: *P* < 0.001 for each meta-analysis), we conducted risk-of-bias sensitivity analyses; meta-analyses of low-risk-of-bias studies continued to show significant heterogeneity (*SI Appendix*, Figs. S3 to S7).

**Fig. 3.**
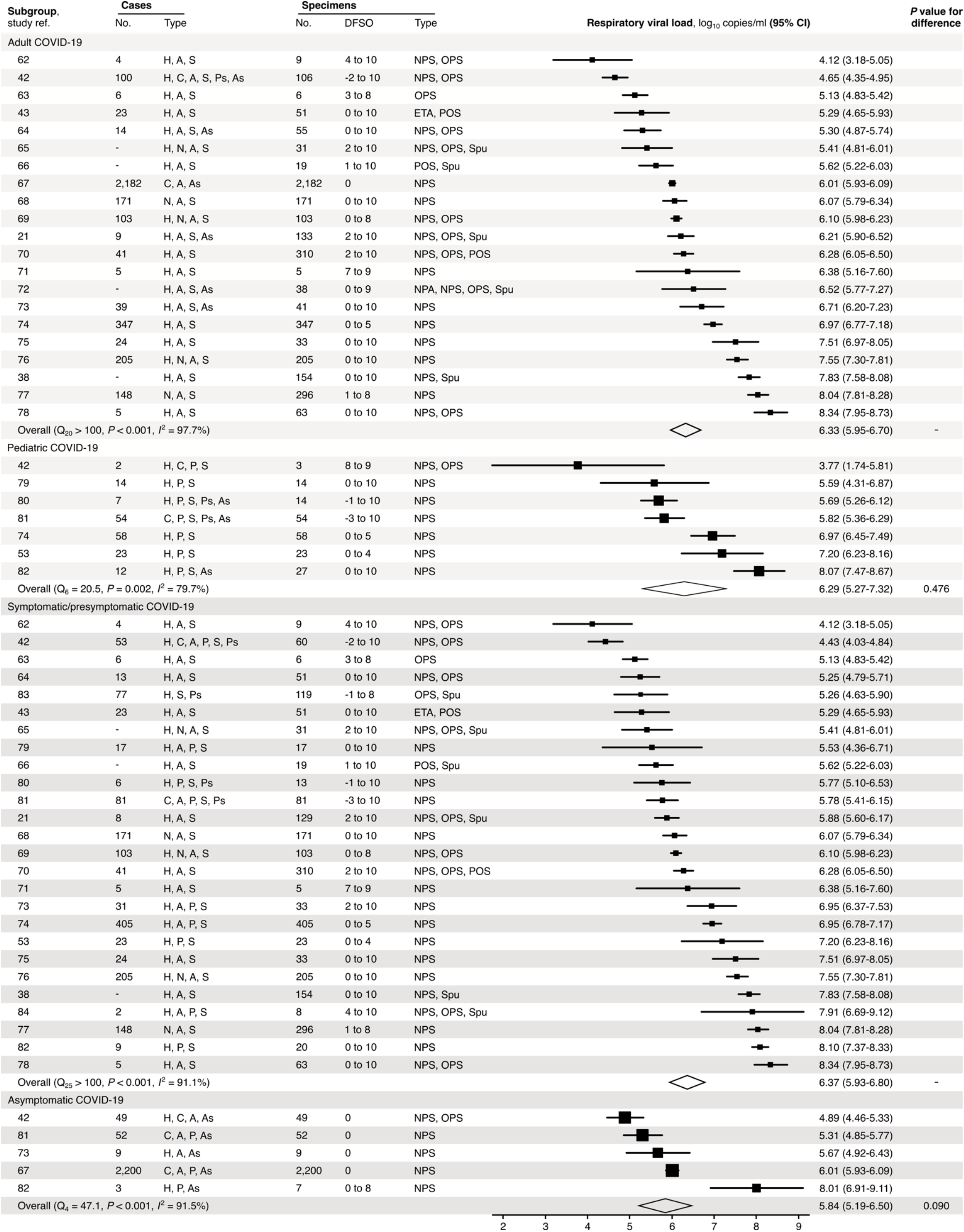
Subgroup analyses of SARS-CoV-2 rVL during the infectious period. Random-effects meta-analyses comparing the expected respiratory viral loads (rVLs) of adult (≥18 years old) COVID-19 cases with pediatric (<18 years old) ones (top) and symptomatic/presymptomatic infections with asymptomatic ones (bottom) during the infectious period. Quantitative rVLs refer to virus concentrations in the respiratory tract. Case types: hospitalized (H), not admitted (N), community (C), adult (A), pediatric (P), symptomatic (S), presymptomatic (Ps) and asymptomatic (As). Specimen types: endotracheal aspirate (ETA), nasopharyngeal aspirate (NPA), nasopharyngeal swab (NPS), oropharyngeal swab (OPS), posterior oropharyngeal saliva (POS) and sputum (Spu). Dashes denote case numbers that were not obtained. Box sizes denote weighting in the overall estimates. Between-study heterogeneity was assessed using the *P*-value from Cochran’s *Q* test and the *I*^2^ statistic. References after 57 are listed in *SI Appendix*, References. One-sided Welch’s *t*-tests compared expected rVLs between the COVID-19 subgroups (non-significance, *P* > 0.05).

### Distributions of rVL

We next analyzed rVL distributions. For all three viruses, rVLs best conformed to Weibull distributions (*SI Appendix*, Fig. S8), and we fitted the entirety of individual sample data for each virus in the systematic dataset (Fig. 4*A* and *SI Appendix*, Fig. S8*N*). While COVID-19 and SARS cases tended to shed lesser virus than those with A(H1N1)pdm09 (*SI Appendix*, Fig. S2), broad heterogeneity in SARS-CoV-2 and SARS-CoV-1 rVLs inverted this relationship for highly infectious individuals (Fig. 4*A* and *SI Appendix*, Fig. S9 *A* to *C*). At the 90^th^ case percentile (cp) throughout the infectious period, the estimated rVL was 8.91 (95% CI: 8.83-9.00) log_10_ copies/ml for SARS-CoV-2, whereas it was 8.62 (8.47-8.76) log_10_ copies/ml for A(H1N1)pdm09 (*SI Appendix*, Table S7). The SD of the overall rVL distribution for SARS-CoV-2 was 2.04 log_10_ copies/ml, while it was 1.45 log_10_ copies/ml for A(H1N1)pdm09, showing that heterogeneity in rVL was indeed broader for SARS-CoV-2.

**Fig. 4.**
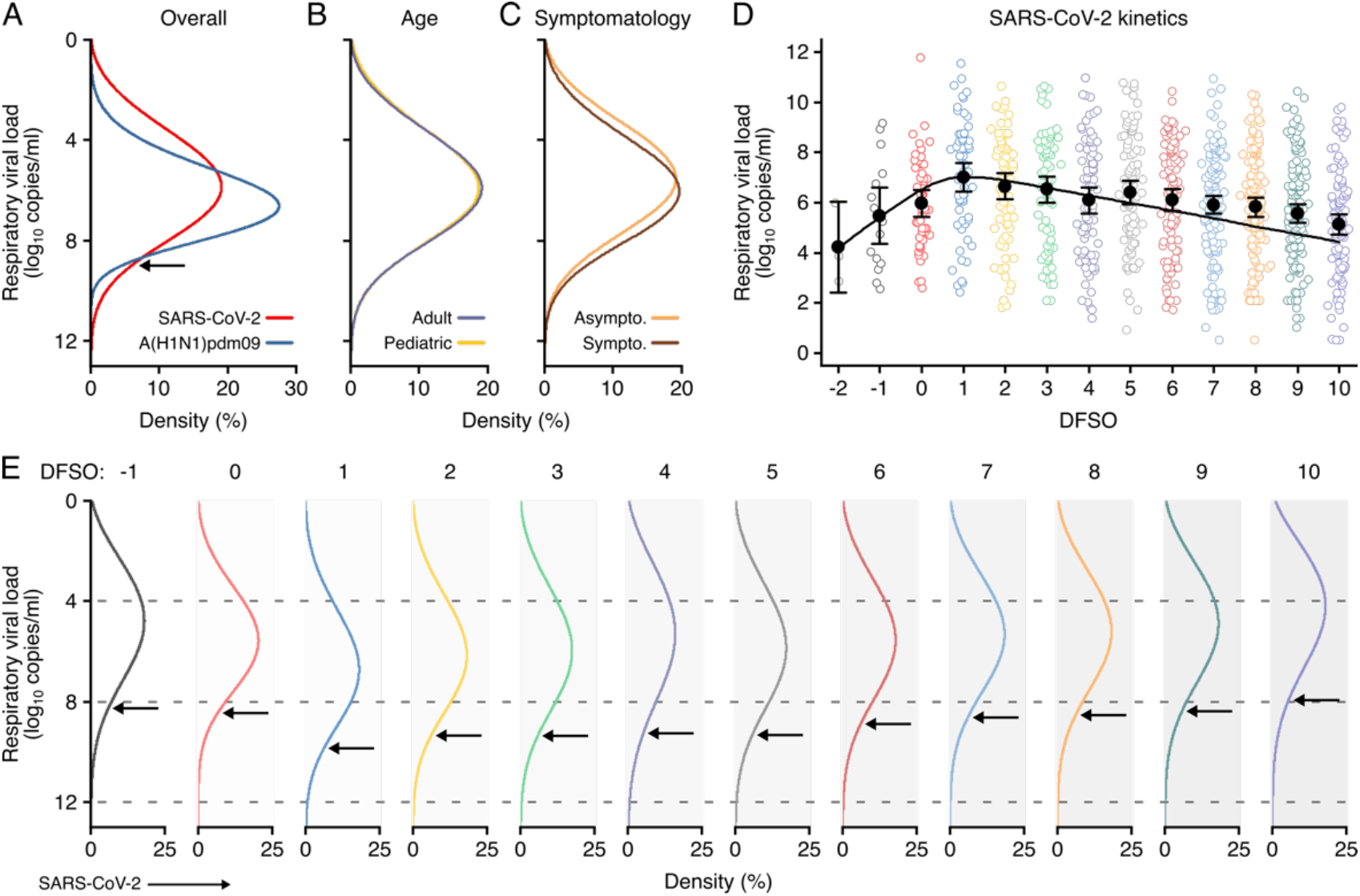
Heterogeneity and kinetics of SARS-CoV-2 rVL. (*A*) Estimated distribution of respiratory viral load (rVL) for SARS-CoV-2 (*N* = 3,834 samples from *N* = 26 studies) and A(H1N1)pdm09 (*N* = 512 samples from *N* = 10 studies) throughout the infectious periods. (*B* and *C*) Estimated distribution of SARS-CoV-2 rVL for adult (*N* = 3,575 samples from *N* = 20 studies) and pediatric (*N* = 198 samples from *N* = 9 studies) (*B*) and symptomatic/presymptomatic (*N* = 1,574 samples from *N* = 22 studies) and asymptomatic (*N* = 2,221 samples from *N* = 7 studies) (*C*) COVID-19 cases. (*D*) SARS-CoV-2 rVLs fitted to a mechanistic model of viral kinetics (black curve, *r*^2^ = 0.84 for mean estimates). Filled circles and bars depict mean estimates and 95% CIs. Open circles show the entirety of individual sample data over days from symptom onset (DFSO) (left to right, *N* = 3, 15, 50, 63, 71, 75, 85, 93, 105, 136, 123, 128 and 115 samples from *N* = 21 studies). (*E*) Estimated distributions of SARS-CoV-2 rVL across DFSO. Weibull distributions were fitted on the entirety of individual sample data for the virus, subgroup or DFSO in the systematic dataset. Arrows denote 90^th^ case percentiles for SARS-CoV-2 rVL distributions.

To assess potential sources of heterogeneity in SARS-CoV-2 rVL, we compared rVL distributions among COVID-19 subgroups. In addition to comparable mean estimates (Fig. 3), adult, pediatric, symptomatic/presymptomatic and asymptomatic COVID-19 cases showed similar rVL distributions (Fig. 4 *B* and *C*), with SDs of 2.03, 2.06, 2.00 and 2.01 log_10_ copies/ml, respectively. Thus, age and symptomatology minimally influenced case variation in SARS-CoV-2 rVL during the infectious period.

### SARS-CoV-2 kinetics during respiratory infection

To analyze the influences of disease course, we delineated individual SARS-CoV-2 rVLs by DFSO and fitted the mean estimates to a mechanistic model for respiratory virus kinetics (Fig. 4*D* and Methods). The outputs indicated that, on average, each productively infected cell in the airway epithelium shed SARS-CoV-2 at 1.33 (95% CI: 0.74-1.93) copies/ml day^-1^ and infected up to 9.25 susceptible cells (*SI Appendix*, Table S8). The turnover rate for infected epithelial cells was 0.71 (0.26-1.15) days^-1^, while the half-life of SARS-CoV-2 in the respiratory tract was 5.04 (2.62-66.0) hours. By extrapolating the model to an initial rVL of 0 log_10_ copies/ml, the estimated incubation period was 5.38 days, which agrees with epidemiological findings (10). Conversely, the expected duration of shedding was 25.1 DFSO. Thus, SARS-CoV-2 rVL increased exponentially after infection, peaked around 1 DFSO along with the proportion of infected epithelial cells (*SI Appendix*, Fig. S10) and then diminished exponentially.

To evaluate case heterogeneity across the infectious period, we fitted distributions for each DFSO (Fig. 4*E*), which showed that high SARS-CoV-2 rVLs also increased from the presymptomatic period, peaked at 1 DFSO and then decreased towards the end of the first week of illness. For the 90^th^ cp at 1 DFSO, the rVL was 9.84 (95% CI: 9.17-10.56) log_10_ copies/ml, an order of magnitude greater than the overall 90^th^-cp estimate. High rVLs between 1-5 DFSO were elevated above the expected values from the overall rVL distribution (*SI Appendix*, Table S7). At -1 DFSO, the 90^th^-cp rVL was 8.30 (6.88-10.02) log_10_ copies/ml, while it was 7.93 (7.35-8.56) log_10_ copies/ml at 10 DFSO. Moreover, heterogeneity in rVL remained broad across the infectious period, with SDs of 1.83-2.44 log_10_ copies/ml between -1 to 10 DFSO (*SI Appendix*, Fig. S9 *H* to *S*).

### Likelihood that droplets and aerosols contain virions

Towards analyzing the influence of heterogeneity in rVL on individual infectiousness, we first modelled the likelihood of respiratory particles containing viable SARS-CoV-2. Since rVL is an intensive quantity, the volume fraction of virions is low and viral partitioning coincides with atomization, we used Poisson statistics to model likelihood profiles. To calculate an unbiased estimator of partitioning (the expected number of viable copies per particle), our method multiplied rVL estimates with particle volumes during atomization and an assumed viability proportion of 0.1% in equilibrated particles (Methods).

When expelled by the mean COVID-19 case during the infectious period, respiratory particles showed low likelihoods of carrying viable SARS-CoV-2 (*SI Appendix*, Fig. S11*B*). Aerosols (equilibrium aerodynamic diameter [*d*_a_] ≤ 100 µm) were ≤0.69% (95% CI: 0.43-0.95%) likely to contain a virion. Droplets also had low likelihoods: at *d*_a_ = 330 µm, they were 19.4% (18.5-20.3%), 2.42% (2.15-2.69%) and 0.20% (0.16-0.24%) likely to contain one, two or three virions, respectively.

COVID-19 cases with high rVLs, however, expelled particles with considerably greater likelihoods of carrying viable copies (Fig. 5 *A* and *B* and *SI Appendix*, Fig. S11 *D* and *E*). For the 80^th^ cp during the infectious period, aerosols (*d*_a_ ≤ 100 µm) were ≤36.6% (95% CI: 35.3-38.0%) likely to carry at least one SARS-CoV-2 virion. For the 90^th^ cp, this likelihood was ≤96.7% (96.5-96.9%), with larger aerosols tending to contain multiple virions (*SI Appendix*, Fig. S11*E*). At 1 DFSO, these estimates were greatest, and ≤61.1% (51.8-70.4%) of buoyant aerosols (*d*_a_ ≤ 10 µm) contained at least one viable copy of SARS-CoV-2 for the 98^th^ cp. When expelled by high cps, droplets (*d*_a_ > 100 µm) tended to contain tens to thousands of SARS-CoV-2 virions (Fig. 5*B* and *SI Appendix*, Fig. S11*E*).

**Fig. 5.**
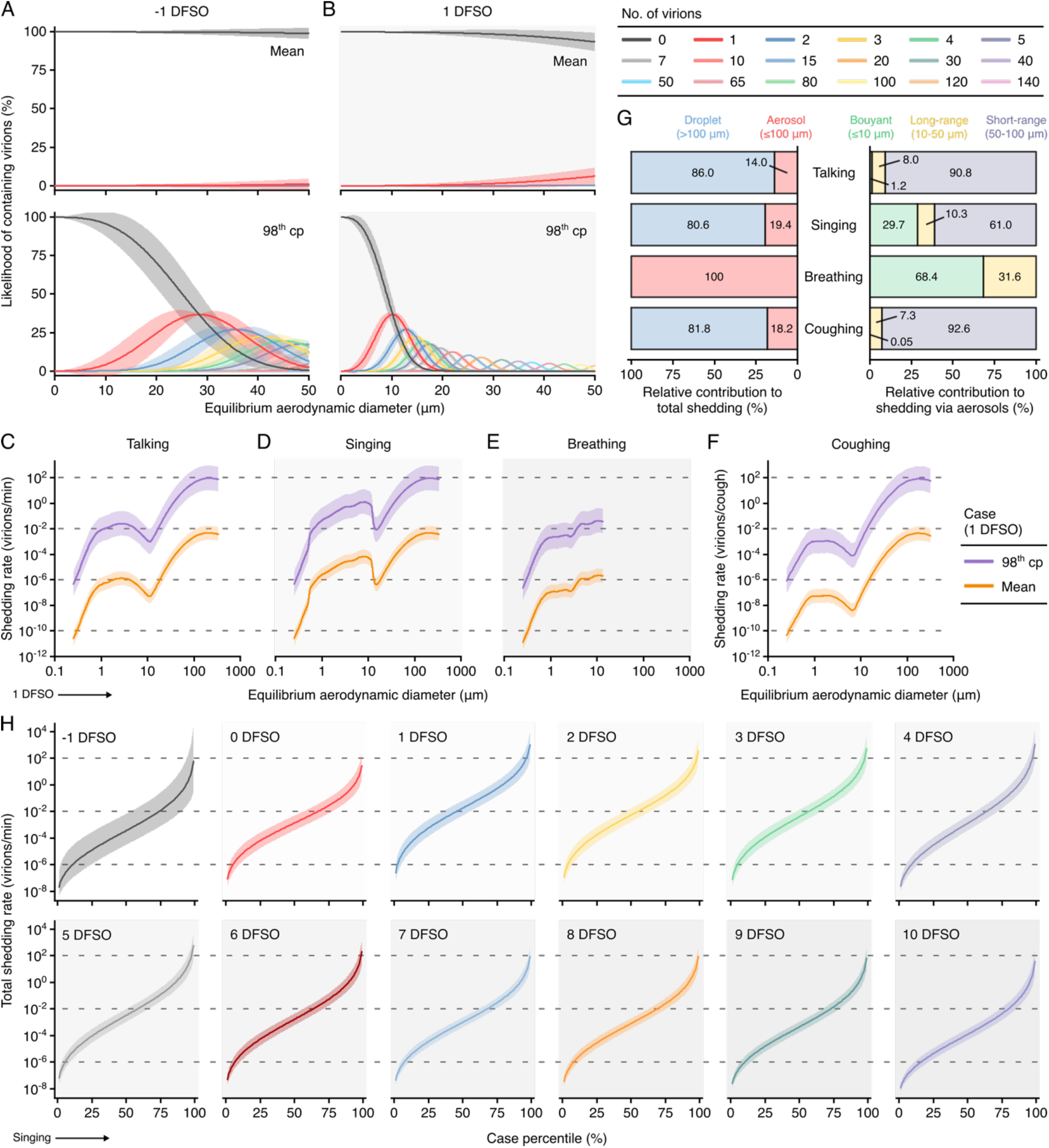
Heterogeneity in shedding SARS-CoV-2 via droplets and aerosols. (*A* and *B*) Estimated likelihood of respiratory particles containing viable SARS-CoV-2 when expelled by the mean (top) or 98^th^ case percentile (cp) (bottom) COVID-19 cases at -1 (*A*) or 1 (*B*) days from symptom onset (DFSO). For higher no. of virions, some likelihood curves were omitted to aid visualization. When the likelihood for 0 virions approaches 0%, particles are expected to contain at least one viable copy. (*C* to *F*) Rate that the mean and 98^th^-cp COVID-19 cases at 1 DFSO shed viable SARS-CoV-2 by talking, singing, breathing or coughing over particle size. (*G*) Relative contributions of droplets and aerosols to shedding virions for each respiratory activity (left). Relative contribution of buoyant, long-range and short-range aerosols to shedding virions via aerosols for each respiratory activity (right). (*H*) Case heterogeneity in the total shedding rate (over all particle sizes) of virions via singing across the infectious period. Earlier presymptomatic days were excluded based on limited data. Data range between the 1^st^ and 99^th^ cps. Lines and bands represent estimates and 95% CIs, respectively, for estimated likelihoods or Poisson means.

### Shedding SARS-CoV-2 via respiratory droplets and aerosols

Using the partitioning estimates in conjunction with published profiles of the particles expelled by respiratory activities (*SI Appendix*, Fig. S12), we next modelled the rates at which talking, singing, breathing and coughing shed viable SARS-CoV-2 across *d*_a_ (Fig. 5 *C* to *F*). Singing shed virions more rapidly than talking based on the increased emission of aerosols. Voice amplitude, however, had a significant effect on aerosol production, and talking loudly emitted aerosols at similar rates to singing (*SI Appendix*, Fig. S12*E*). Based on the generation of larger aerosols and droplets, talking and singing shed virions more rapidly than breathing (Fig. 5 *C* to *E*). Each cough shed similar quantities of virions as in a minute of talking (Fig. 5 *C* and *F*).

Each of these respiratory activities expelled aerosols at greater rates than droplets, but particle size correlated with the likelihood of containing virions. Talking, singing and coughing expelled virions at greater proportions via droplets (80.6-86.0%) than aerosols (14.0-19.4%) (Fig. 5*G*). Moreover, short-range aerosols predominantly mediated the virions (90.8-92.6%) shed via aerosols while talking normally and coughing. In comparison, while singing, talking loudly and breathing, buoyant (29.7-68.4%) and long-range (10.3-31.6%) aerosols carried a larger proportion of the virions shed via aerosols (Fig. 5*G*).

### Influence of heterogeneity in rVL on individual infectiousness

To interpret how heterogeneity in rVL influences individual infectiousness, we modelled total SARS-CoV-2 shedding rates (over all particle sizes) for each respiratory activity (Fig. 5*H* and *SI Appendix*, Fig. S13). Between the 1^st^ and the 99^th^ cps, the estimates for a respiratory activity spanned ≥8.48 orders of magnitude on each DFSO; cumulatively from -1 to 10 DFSO, they spanned 11.0 orders of magnitude. Hence, many COVID-19 cases inherently presented minimal transmission risk, whereas highly infectious individuals shed considerable quantities of SARS-CoV-2. For the 98^th^ cp at 1 DFSO, singing expelled 313 (95% CI: 37.5-3,158) virions/min to the ambient environment, talking emitted 293 (35.1-2,664) virions/min, breathing exhaled 1.54 (0.18-15.5) virions/min and coughing discharged 249 (29.8-2,5111) virions/cough; these estimates were approximately two orders of magnitude greater than those for the 85^th^ cp. For the 98^th^ cp at -1 DFSO, singing shed 14.5 (0.15-4,515) virions/min and breathing exhaled 7.13×10^−2^ (7.20×10^−4^-22.2) virions/min. The estimates at 9-10 DFSO were similar to these presymptomatic ones (Fig. 5*H* and *SI Appendix*, Fig. S13*B*). As indicated by comparable mean rVLs (Fig. 3) and heterogeneities in rVL (Fig. 4 *B* and *C*), adult, pediatric, symptomatic/presymptomatic and asymptomatic COVID-19 subgroups presented similar distributions for shedding virions through these activities.

We also compared the influence of case variation on individual infectiousness between A(H1N1)pdm09 and COVID-19. Aerosol spread accounted for approximately half of A(H1N1)pdm09 transmission events (19), and the 50% human infectious dose for aerosolized influenza A virus is approximately 1-3 virions in the absence of neutralizing antibodies (26). Based on the model, 62.9% of A(H1N1)pdm09 cases were infectious (shed ≥1 virion) via aerosols within 24 h of talking loudly or singing (*SI Appendix*, Fig. S14*A*). The estimate was 58.6% within 24 h of talking normally and 22.3% within 24 h of breathing. In comparison, 48.0% of COVID-19 cases shed ≥1 virion via aerosols in 24 h of talking loudly or singing (*SI Appendix*, Fig. S14*C*). Notably, only 61.4% of COVID-19 cases shed ≥1 virion via either droplets or aerosols in 24 h of talking loudly or singing (*SI Appendix*, Fig. S14*D*). While the human infectious dose of SARS-CoV-2 by any exposure route remains unelucidated, it must be at least one viable copy. Thus, at least 38.6% of COVID-19 cases were expected to present negligible risk to spread SARS-CoV-2 through either droplets or aerosols in 24 h. The proportion of inherently infectious cases further decreased as the infectious dose increased: 55.8, 42.5 and 25.0% of COVID-19 cases were expected to shed ≥2, ≥10 and ≥100 virions, respectively, in 24 h of talking loudly or singing during the infectious period.

While these analyses indicated that a greater proportion of A(H1N1)pdm09 cases were inherently infectious, 18.8% of COVID-19 cases shed virions more rapidly than those infected with A(H1N1)pdm09 (Fig. 4*A*). At the 98^th^ cp for A(H1N1)pdm09, singing expelled 4.38 (2.85-6.78) virions/min and breathing exhaled 2.15×10^−2^ (1.40×10^−2^-3.34×10^−2^) virions/min. Highly infectious COVID-19 cases expelled virions at rates that were up to 1-2 orders of magnitude greater than their A(H1N1)pdm09 counterparts (Fig. 5*H* and *SI Appendix*, Fig. S15).

## Discussion

This study provided systematic analyses of several factors characterizing SARS-CoV-2 transmissibility. First, we found that broader heterogeneity in rVL facilitates greater overdispersion for SARS-CoV-2 than A(H1N1)pdm09. Our results suggest that many COVID-19 cases infect no one (12-14) because they inherently present minimal transmission risk via droplets or aerosols, although behavioral and environmental factors can further abate risk. Meanwhile, highly infectious cases can shed tens to thousands of SARS-CoV-2 virions/min, especially between 1-5 DFSO. The model estimates, when corrected to copies rather than virions, align with recent clinical findings for exhalation rates of SARS-CoV-2 (27). In comparison, a greater proportion of A(H1N1)pdm09 cases are infectious but shed virions at low rates, which concurs with more uniform transmission and few superspreading events observed during the 2009 H1N1 pandemic (16, 17). Moreover, our analyses suggest that heterogeneity in rVL may be generally associated with overdispersion for viral respiratory infections. In this case, rVL distribution can serve as an early correlate for transmission patterns, including superspreading, during outbreaks of novel respiratory viruses, providing insight for disease control before large-scale epidemiological studies empirically characterize *k*. When transmission is highly overdispersed, targeted interventions may disproportionately mitigate infection (20), with models estimating that focusing half of control efforts on the most infectious 20% of cases outperforms random control policies threefold (15).

Second, we analyzed SARS-CoV-2 kinetics during respiratory infection. While heterogeneity remains broad throughout the infectious period, rVL tends to peak at 1 DFSO and be elevated for 1-5 DFSO, coinciding with the period of highest attack rates observed among close contacts (28). These results indicate that transmission risk tends to be greatest soon after illness rather than in the presymptomatic period, which concurs with large tracing studies (6.4-12.6% of secondary infections from presymptomatic transmission) (29, 30) rather than early temporal models (∼44%) (23). Furthermore, our kinetic analysis suggests that, on average, SARS-CoV-2 reaches diagnostic concentrations 1.54-3.17 days after respiratory infection (−3.84 to -2.21 DFSO), assuming assay detection limits of 1-3 log_10_ copies/ml, respectively, for nasopharyngeal swabs immersed in 1 ml of transport media.

Third, we assessed the relative infectiousness of COVID-19 subgroups. As a common symptom of COVID-19 (31), coughing sheds considerable numbers of virions via droplets and short-range aerosols. Thus, symptomatic infections tend to be more contagious than asymptomatic ones, providing one reason as to why asymptomatic cases transmit SARS-CoV-2 at lower relative rates (32), especially in close contact (33), despite similar rVLs and increased contact patterns. Accordingly, children (48-54% of symptomatic cases present with cough) (34, 35) tend to be less contagious than adults (68-80%) (31, 35) based on tendencies of symptomatology rather than rVL. Conversely, coughing sheds few virions via smaller aerosols. Our analyses suggest that asymptomatic and symptomatic infections present comparable risks for airborne spread, as do adult and pediatric cases. While singing and talking loudly, highly infectious cases shed tens to hundreds of SARS-CoV-2 virions/min via long-range and buoyant aerosols.

Our study has limitations. The systematic search found a limited number of studies reporting quantitative specimen measurements from the presymptomatic period, meaning these estimates may be sensitive to sampling bias. Although additional studies have reported semiquantitative metrics (cycle thresholds), these data were excluded because they cannot be compared on an absolute scale due to batch effects (36), limiting use in compound analyses. Furthermore, this study considered population-level estimates of the infectious periods, viability proportions and rate profiles for respiratory particles, which omit individual or environmental variation. Distinctions in phonetic tendencies and, especially for young children, respiratory capacity lead to variation in particle emission rates (37). Some patients shed SARS-CoV-2 with diminishing viability soon after symptom onset (21), whereas others produce replication-competent virus for weeks (38). It remains unclear how case characteristics and environmental factors affect the viability dynamics of SARS-CoV-2.

Taken together, our findings provide a potential path forward for disease control. They support aerosol spread as a transmission mode for SARS-CoV-2, including for conditional superspreading by highly infectious cases. However, with short durations of stay in well-ventilated areas, the exposure risk for aerosols, including long-range and buoyant ones, remains correlated with proximity to infectious cases (2, 4). Strategies to abate infection should limit crowd numbers and duration of stay while reinforcing distancing, low voice amplitudes and widespread mask usage; well-ventilated settings can be recognized as lower risk venues. Coughing can shed considerable quantities of virions, while rVL tends to peak at 1 DFSO and can be high throughout the infectious period. Thus, immediate, sustained self-isolation upon illness is crucial to curb transmission from symptomatic cases. Collectively, our analyses highlight the role of cases with high rVLs in propelling the COVID-19 pandemic. While diagnosing COVID-19, qRT-PCR can also triage contact tracing, prioritizing these patients: for nasopharyngeal swabs immersed in 1 ml of transport media, ≥7.14 (95% CI: 7.07-7.22) log_10_ copies/ml corresponds to the top 20% of COVID-19 cases. Doing so may identify asymptomatic and presymptomatic infections more efficiently, a key step towards mitigation as the pandemic continues.

## Materials and Methods

### Systematic review

We undertook a systematic review and prospectively submitted the protocol for registration on PROSPERO (registration number, CRD42020204637). Other than the title of this study, we have followed PRISMA reporting guidelines (39). The systematic review was conducted according to Cochrane methods guidance (40). In-depth details on the search strategy, selection criteria, data collection and contributing studies are included in the *SI Appendix*, Methods.

### Calculation of rVLs from specimen measurements

In this study, viral concentrations in respiratory specimens were denoted as specimen measurements, whereas viral concentrations in the respiratory tract were denoted as rVLs. To determine rVLs, each collected quantitative specimen measurement was converted to rVL based on the dilution factor. For example, measurements from swabbed specimens (NPS and OPS) typically report the RNA concentration in viral transport media. Based on the expected uptake volume for swabs (0.128 ± 0.031 ml, mean ± SD) (41) or reported collection volume for expulsed fluid in the study (e.g., 0.5 to 1 ml) along with the reported volume of transport media in the study (e.g., 1 ml), we calculated the dilution factor for each respiratory specimen to estimate the rVL. If the diluent volume was not reported, then the dilution factor was calculated assuming a volume of 1 ml (NPS and OPS), 2 ml (POS and ETA) or 3 ml (NPA) of transport media (42-44). Unless dilution was reported for Spu specimens, we used the specimen measurement as the rVL (21). The non-reporting of diluent volume was noted as an element increasing risk of bias in the hybrid JBI critical appraisal checklist. Specimen measurements (based on instrumentation, calibration, procedures and reagents) are not standardized. While the above procedures (including only quantitative measurements after extraction as an inclusion criterion, considering assay detection limits and correcting for specimen dilution) have considered many of these factors, non-standardization remains an inherent limitation in the variability of specimen measurements.

### Meta-regression of *k* and heterogeneity in rVL

To assess the relationship between *k* and heterogeneity in rVL, we performed a univariate meta-regression (log *k* = *a*(*SD*) + *b*, where *a* is the slope for association and *b* is the intercept) between pooled estimates of *k* (based on studies describing community transmission) for COVID-19 (*k* = 0.409) (12-14, 45-48), SARS (*k* = 0.165) (15) and A(H1N1)pdm09 (*k* = 8.155) (16, 17) and the SD of the rVLs in contributing studies. Since SD was the metric, we used a fixed-effects model. For weighting in the meta-regression, we used the proportion of rVL samples from each study relative to the entire systematic dataset (*W*_i_ = *n*_i_/*n*_total_). All calculations were performed in units of log_10_ copies/ml. As the meta-regression used pooled estimates of *k* for each infection, it assumed that there was no correlated bias to *k* across contributing studies. The limit of detection for qRT-PCR instruments used in the included studies did not significantly affect the analysis of heterogeneity in rVL, as these limits tended to be below the values found for specimens with low virus concentrations. The meta-regression was conducted using all contributing studies and showed a weak association. Meta-regression was also conducted using studies that had low risk of bias according to the hybrid JBI critical appraisal checklist and showed a strong association. The *P*-value for association was obtained using the meta-regression slope *t*-test for *a*, the effect estimate.

### Meta-analysis of rVLs

Based on the search design and composition of contributing studies, the meta-analysis overall estimates were the expected SARS-CoV-2, SARS-CoV-1 and A(H1N1)pdm09 rVL when encountering a COVID-19, SARS or A(H1N1)pdm09 case, respectively, during their infectious period. Pooled estimates and 95% CIs for the expected rVL of each virus across their infectious period were calculated using a random-effects meta-analysis (DerSimonian and Laird method). The estimates for rVL assumed that each viral copy was extracted and quantified from the tested specimen aliquot. For studies reporting summary statistics in medians and interquartile or total ranges, we derived estimates of the mean and variance and calculated the 95% CIs (49). All calculations were performed in units of log_10_ copies/ml. Between-study heterogeneity in meta-analysis was assessed using Cochran’s *Q* test and the *I*^2^ and τ^2^ statistics. If significant between-study heterogeneity in meta-analyses was encountered, sensitivity analysis based on the risk of bias of contributing studies was performed. The meta-analyses were conducted using STATA 14.2 (StataCorp LLC, College Station, Texas, USA).

### Age and symptomatology subgroup analyses of SARS-CoV-2 rVLs

The overall estimate for each subgroup was the expected rVL when encountering a case of that subgroup during the infectious period. Studies reporting data exclusively from a subgroup of interest were directly included in the analysis after rVL estimations. For studies in which data for these subgroups constituted only part of its dataset, rVLs from the subgroup were extracted to calculate the mean, variance and 95% CIs. Random-effects meta-analysis was performed as described above. For meta-analyses of pediatric and asymptomatic COVID-19 cases, contributing studies had low risk of bias, and no risk-of-bias sensitivity analyses were performed for these subgroups.

### Distributions of rVL

We pooled the entirety of individual sample data in the systematic dataset by disease, COVID-19 subgroups and DFSO. For analyses of SARS-CoV-2 dynamics across disease course, we included estimated rVLs from negative qRT-PCR measurements of respiratory specimens for cases that had previously been quantitatively confirmed to have COVID-19. These rVLs were estimated based on the reported assay detection limit in the respective study. Probability plots and modified Kolmogorov–Smirnov tests used the Blom scoring method and were used to determine the suitability of normal, lognormal, gamma and Weibull distributions to describe the distribution of rVLs for SARS-CoV-2, SARS-CoV-1 and A(H1N1)pdm09. For each virus, the data best conformed to Weibull distributions, which is described by the probability density function

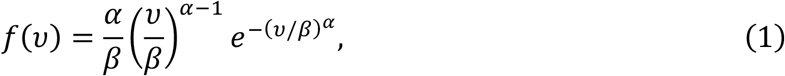

where *α* is the shape factor, *β* is the scale factor and *ν* is rVL (*ν* ≥ 0 log_10_ copies/ml). Weibull distributions were fitted on the entirety of collected individual sample data for the respective category. Since individual specimen measurements could not be collected from all studies, there was a small bias on the mean estimate for each fitted distribution. Thus, for the curves shown in Fig. 4, *B* and *C*, the mean of the Weibull distributions summarized in *SI Appendix*, Table S7 was adjusted to be the subgroup meta-analysis estimate for correction; the SD and distribution around that mean remained consistent.

For each Weibull distribution, the value of the rVL at the *x*^th^ percentile was determined using the quantile function,

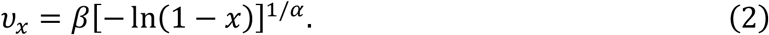

For cp curves, we used eq. (2) to determine rVLs from the 1^st^ cp to the 99^th^ cp (step size, 1%). Curve fitting to eq. (1) and calculation of eq. (2) and its 95% CI was performed using the Distribution Fitter application in Matlab R2019b (MathWorks, Inc., Natick, Massachusetts, USA).

### Viral kinetics

To model SARS-CoV-2 kinetics during respiratory infection, we used a mechanistic epithelial cell-limited model for the respiratory tract (50), based on the system of differential equations:

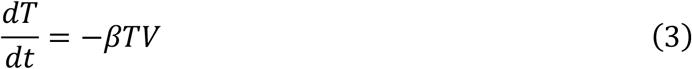

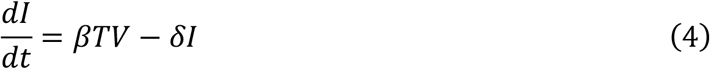

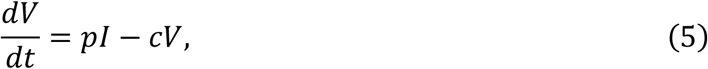

where *T* is the number of uninfected target cells, *I* is the number of productively infected cells, *V* is the rVL, *β* is the infection rate constant, *p* is the rate at which airway epithelial cells shed virus to the extracellular fluid, *c* is the clearance rate of virus and *δ* is the clearance rate of productively infected cells. Parameter units are summarized in *SI Appendix*, Table S8. Using these parameters, the viral half-life in the respiratory tract (*t*_1/2_ = ln 2/*c*) and the half-life of productively infected cells (*t*_1/2_ = ln 2/*δ*) could be estimated. Moreover, the cellular basic reproductive number (the expected number of secondary infected cells from a single productively infected cell placed in a population of susceptible cells) was calculated by

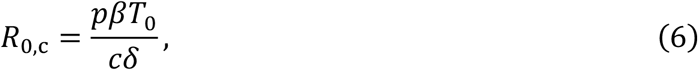

where *T*_0_ is the initial number of susceptible cells (50). Further details on fitting procedure and analyses of viral kinetics are included in the *SI Appendix*, Methods.

### Likelihood of respiratory particles containing virions

To calculate an unbiased estimator for viral partitioning (the expected number of viable copies in an expelled particle at a given size), we multiplied rVLs with the volume equation for spherical particles during atomization and the estimated viability proportion, according to the following equation:

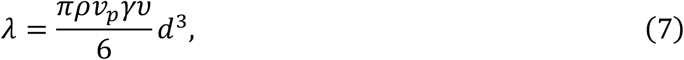

where *λ* is the expectation value, *ρ* is the material density of the respiratory particle (997 g/m^3^), *υ*_*p*_ is the volumetric conversion factor (1 ml/g), *γ* is the viability proportion, *ν* is the rVL and *d* is the hydrated diameter of the particle during atomization.

The model assumed *γ* was 0.1% as a population-level estimate. For influenza, approximately 0.1% of copies in particles expelled from the respiratory tract represent viable virus (51), which is equivalent to one in 3 log_10_ copies/ml for rVL or, after dilution in transport media, roughly one in 4 log_10_ copies/ml for specimen concentration. Respiratory specimens taken from influenza cases show positive cultures for specimen concentrations down to 4 log_10_ copies/ml (52). Likewise, for COVID-19 cases, recent reports also show culture-positive respiratory specimens with SARS-CoV-2 concentrations down to 4 log_10_ copies/ml (21), including from pediatric (53) and asymptomatic (22) cases. Our analyses indicated that SARS-CoV-2 rVLs were not different at different sites in the respiratory tract. Moreover, replication-competent SARS-CoV-2 has been found in respiratory specimens taken throughout the respiratory tract (mouth, nasopharynx, oropharynx and lower respiratory tract) (21, 54). Taken together, these considerations suggested that the assumption for viability proportion (0.1%) was suitable to model the likelihood of respiratory particles containing viable SARS-CoV-2. In accordance with the discussion above, the model did not differentiate this population-level viability estimate based on age, symptomatology or sites of atomization. Based on the relative relationship between the residence time of expelled particles before assessment (∼5 s) (51), we took the viability proportion (0.1%) to be for equilibrated particles.

Likelihood profiles were determined using Poisson statistics, as described by the probability mass function

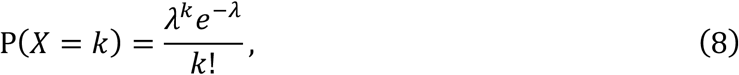

where *k* is the number of virions partitioned within the particle. For *λ*, 95% CIs were determined using the variance of its rVL estimate. To determine 95% CIs for likelihood profiles from the probability mass function, we used the delta method, which specifies

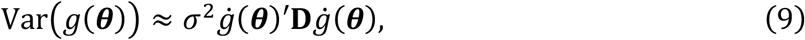

where *σ*^2^**D** is the covariance matrix of ***θ*** and 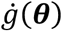 is the gradient of *g*(***θ***). For the univariate Poisson distribution, *σ*^2^**D** = *λ* and

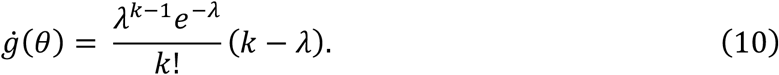

### Rate profiles of particles expelled by respiratory activities

Distributions from the literature were used to determine the rate profiles of particles expelled during respiratory activities. For breathing, talking and coughing, we used data from Johnson et al (55). For singing, we used data from Morawaska et al (56) for smaller aerosols (*d*_a_ < 20 μm) and used the profiles from talking for larger aerosols and droplets based on the oral cavity mechanism from Johnson et al (55). Further details on the calculation of rate profiles are included in the *SI Appendix*, Methods. We compared these rate profiles with those collected from talking loudly and talking quietly from Asadi et al (57). After dehydration, the particle diameter becomes approximately 0.5 times the initial size from when atomized in the respiratory tract (55), which was used as the factor between the diameter of hydrated particles during atomization and dehydrated particles.

Equilibrium aerodynamic diameter was calculated by *d*_*a*_ = *d*_p_(*ρ*/*ρ*_0_)^1/2^, where *d*_*p*_ is the dehydrated diameter, *ρ* is the material density of the respiratory particle (taken to be 1 g/cm^3^ based on the composition of dehydrated respiratory particles) and *ρ*_0_ is the reference material density (1 g/cm^3^). Curves based on discrete particle measurements were connected using the nonparametric Akima spline function.

### Shedding virions via respiratory droplets and aerosols

To model the respiratory shedding rate across particle size, rVL estimates and the hydrated diameters of particles expelled by a respiratory activity were input into eq. (7), and the output was then multiplied by the rate profile of the activity (talking, singing, breathing or coughing). To assess the relative contribution of aerosols and droplets to mediating respiratory viral shedding for a given respiratory activity, we calculated the proportion of the cumulative hydrated volumetric rate contributed by buoyant aerosols (*d*_a_ ≤ 10 μm), long-range aerosols (10 μm < *d*_a_ ≤ 50 μm), short-range aerosols (50 μm < *d*_a_ ≤ 100 μm) and droplets (*d*_a_ > 10 μm) for that respiratory activity. Since the Poisson mean was proportional to cumulative volumetric rate, this estimate of the relative contribution of aerosols and droplets to respiratory viral shedding was consistent among viruses and cps in the model.

To determine the total respiratory shedding rate for a given respiratory activity across cp, we determined the cumulative hydrated volumetric rate (by summing the hydrated volumetric rates across particle sizes for that respiratory activity) of particle atomization and input it into eq. (7). Using rVLs and their variances as determined by the Weibull quantile functions, we then calculated the Poisson means and their 95% CIs at the different cps.

To assess the influence of heterogeneity in rVL on individual infectiousness, we first considered transmission of A(H1N1)pdm09 via aerosols (19). The 50% human infectious dose (HID_50_) of aerosolized A(H1N1)pdm09 was taken to be 1-3 virions (26). To determine the expected time required for a A(H1N1)pdm09 case to shed 1 virion via aerosols, we took the reciprocal of the Poisson means and their 95% CIs at the different cps of the estimated shedding rates. The expected time required for a COVID-19 case to shed 1 virion via aerosols or 1 virion via droplets or aerosols was determined in a same manner.

## Data Availability

Data will be made available upon request. All raw data, code and model outputs from this study will be made publicly available in online repositories after peer review. Search strategies for the systematic review are shown in Supplementary Tables 1-5. The systematic review protocol was prospectively registered on PROSPERO (registration number, CRD42020204637).

## Acknowledgments

We thank T. Alba (Toronto) for discussion on statistical methods. We thank J. Jimenez (Colorado) for discussion on the characteristics of aerosols and droplets. We thank E. Lavezzo and A. Chrisanti (Padova) and A. Wyllie, A. Ko and N. Grubaugh (Yale) for responses to data inquiries. This work was supported by the Natural Sciences and Engineering Research Council of Canada (NSERC) and the Toronto COVID-19 Action Fund. P.Z.C. was supported by the NSERC Vanier Scholarship (608544). D.N.F. was supported by the Canadian Institutes of Health Research (Canadian COVID-19 Rapid Research Fund, OV4-170360). F.X.G. was supported by the NSERC Senior Industrial Research Chair.

## Author Contributions

P.Z.C. designed research, analyzed data and wrote the paper. P.Z.C. and N.B. conducted screening, appraised studies and drafted the review protocol. Z.P. developed and conducted the systematic review search. M.K. and D.N.F. assessed methods, interpreted results and contributed to the discussion. All authors reviewed and revised the manuscript. F.X.G. supervised the research.

