## Supplementary material for "Heterogeneity in transmissibility and shedding SARS-CoV-2 via droplets and aerosols": SI Appendix

**This PDF file includes:**

Supplementary Methods
Figures S1 to S15
Tables S1 to S10
SI References

### Supplementary Methods

**Search strategy, selection criteria and data collection.** The search included papers that (i) reported positive, quantitative measurements (copies/ml or an equivalent metric) of SARS-CoV-2, SARS-CoV-1 or A(H1N1)pdm09 in human respiratory specimens (ETA, NPA, NPS, OPS, POS and Spu) from COVID-19, SARS or A(H1N1)pdm09 cases; (ii) reported data that could be extracted from the infectious periods of SARS-CoV-2 (defined as -3 to +10 DFSO for symptomatic cases and 0 to +10 days from the day of laboratory diagnosis for asymptomatic cases), SARS-CoV-1 (defined as 0 to +20 DFSO or the equivalent asymptomatic period) or A(H1N1)pdm09 (defined as -2 to +9 DFSO for symptomatic cases and 0 days to +9 days from the day of laboratory diagnosis for asymptomatic cases); and (iii) reported data for two or more cases with laboratory-confirmed COVID-19, SARS or A(H1N1)pdm09 based on World Health Organization (WHO) case definitions. Quantitative specimen measurements were considered after RNA extraction for diagnostic sequences of SARS-CoV-2 (*Ofr1b*, *N*, *RdRp* and *E* genes), SARS-CoV-1 (*Ofr1b*, *N* and *RdRp* genes) and A(H1N1)pdm09 (*HA* and *M* genes).

Studies were excluded, in the following order, if they (i) studied an ineligible disease; (ii) had an ineligible study design, including those that were reviews of evidence (e.g., scoping, systematic or narrative), did not include primary clinical human data, reported data for less than two cases due to an increased risk of selection bias, were incomplete (e.g., ongoing clinical trials), did not report an RNA extraction step before measurement or were studies of environmental samples; (iii) reported an ineligible metric for specimen concentration (e.g., qualitative RT-PCR or cycle threshold [Ct] values without calibration included in the study); (iv) reported quantitative measurements from an ineligible specimen type (e.g., blood specimens, pooled specimens or self-collected POS or Spu patient specimens in the absence of a healthcare professional); (v) reported an ineligible sampling period (consisted entirely of data that could not be extracted from within the infectious period); or (vi) were duplicates of an included study (e.g., preprinted version of a published paper or duplicates not identified by Covidence). We included data from control groups receiving standard of care in interventional studies but excluded data from the intervention group. Patients in the intervention group are, by definition, systematically different from general case populations because they receive therapies not being widely used for treatment, which may influence virus concentrations. Interventional studies examining the comparative effectiveness of two or more treatments were excluded for the same reason. Studies exclusively reporting semiquantitative measurements (e.g., Ct values) of specimen concentration were excluded, as these measurements are sensitive to batch and instrument inconsistencies and, without proper calibration, cannot be compared on an absolute scale across studies (36).

We searched, without the use of filters or language restrictions, the following sources: MEDLINE (via Ovid, 1946 to 7 August 2020), EMBASE (via Ovid, 1974 to 7 August 2020, Cochrane Central Register of Controlled Trials (via Ovid, 1991 to 7 August 2020), Web of Science Core Collection (including: Science Citation Index Expanded, 1900 to 7 August 2020; Social Sciences Citation Index, 1900 to 7 August 2020; Arts & Humanities Citation Index, 1975 to 7 August 2020; Conference Proceedings Citation Index - Science, 1990 to 7 August 2020; Conference Proceedings Citation Index - Social Sciences & Humanities, 1990 to 7 August 2020; and Emerging Sources Citation Index, 2015 to 7 August 2020), as well as medRxiv and bioRxiv (both searched through Google Scholar via the Publish or Perish program, to 7 August 2020). We also gathered studies by searching through the reference lists of review articles identified by the database search, by searching through the reference lists of included articles, through expert recommendation (by Epic J. Topol, Akiko Iwasaki and A. Marm Kilpatrick on Twitter) and by hand-searching through journals (*Nature*, *Nat. Med.*, *Science*, *NEJM*, *Lancet*, *Lancet Infect. Dis.*, *JAMA*, *JAMA Intern. Med.* and *BMJ*). A comprehensive search was developed by a librarian, which included subject headings and keywords. The search strategy had 3 main concepts (disease, specimen type and outcome), and each concept was combined using the appropriate Boolean operators. The search was tested against a sample set of known articles that were pre-identified. The line-by-line search strategies for all databases are included in *SI Appendix*, Tables S1 to S5. The search results were exported from each database and uploaded to the Covidence online system for deduplication and screening.

Two authors independently screened titles and abstracts, reviewed full texts, collected data and assessed risk of bias via Covidence and a hybrid critical appraisal checklist based on the Joanna Briggs Institute (JBI) tools for case series, analytical cross-sectional studies and prevalence studies (58-60). To evaluate the sample size in a study, we used the following calculation:

$$n^* = \frac{z^2\sigma}{d^2}, \tag{S1}$$

where $n^*$ is the sample size threshold, $z$ is the z-score for the level of confidence (95%), $\sigma$ is the standard deviation (assumed to be 3 $\log_{10}$ copies/ml, one quarter of the full range of rVLs) and $d$ is the marginal error (assumed to be 1 $\log_{10}$ copies/ml, based on the minimum detection limit for qRT-PCR across studies) (61). The hybrid JBI critical appraisal checklist is shown in *SI Appendix*, Table S10. Studies were considered to have low risk of bias if they met the majority of the items, indicating that the estimates were likely to be correct for the target population. Inconsistencies were resolved by discussion and consensus.

The search found 29 studies for COVID-19 (21, 38, 42, 43, 53, 62-85), 8 studies for SARS (44, 86-92) and 27 studies for A(H1N1)pdm09 (93-119) and data were collected from each study. For preprinted studies that were published as journal articles before the submission date of this manuscript, we included the citation for the journal article. Descriptive statistics on quantitative specimen measurements were collected from confirmed cases directly if reported numerically or using WebPlotDigitizer 4.3 (https://apps.automeris.io/wpd/) if reported graphically. Individual specimen measurements were collected directly if reported numerically or, when the data were clearly represented, using the tool if reported graphically. We also collected the relevant numbers of cases, types of cases, pharmacotherapies, volumes of transport media, numbers of specimens and DFSO (for symptomatic cases) or day relative to initial laboratory diagnosis (for asymptomatic cases) on which each specimen was taken. Hospitalized cases were defined as those being tested in a hospital setting and then admitted. Non-admitted cases were defined as those being tested in a hospital setting but not admitted. Community cases were defined as those being tested in a community setting. Symptomatic, presymptomatic and asymptomatic infections were defined as in the study. Based on rare description in contributing studies, paucisymptomatic infections, when described, were included with symptomatic ones. Pediatric cases were defined as those below 18 years of age or as defined in the study. Adult cases were defined as those 18 years of age or higher or as defined in the study.

**Viral kinetics.** For initial parameterization, eqs. (4)-(6) were simplified according to a quasi-steady state approximation (120) to

$$\frac{dT}{dt} = -\beta TV \tag{S2}$$

$$\frac{dV}{dt} = rTV - \delta V, \tag{S3}$$

where $r = p\beta/c$, for a form with greater numerical stability. The system of differential equations was fitted on the mean estimates of SARS-CoV-2 rVL between -2 and 10 DFSO using the entirety of individual sample data in units of copies/ml. Numerical analysis was implemented using the Fit ODE app in OriginPro 2019b (OriginLab Corporation, Northampton, Massachusetts, USA) via the Runge-Kutta method and initial parameters $V_0$, $I_0$ and $T_0$ of 4 copies/ml, 0 cells and $5\times10^7$ cells, respectively, for the range -5 to 10 DFSO. The analysis was first performed with eqs. (S2)-(S3). These output parameters were then used to initialize final analysis using eqs. (3)-(5), where the estimates for $\beta$ and $\delta$ were input as fixed and variable parameters, respectively. The fitted line and its coefficient of determination ($r^2$) were presented.

To estimate the average incubation period, we extrapolated the kinetic model to 0 $\log_{10}$ copies/ml pre-symptom onset. To estimate the average duration of shedding, we extrapolated the model to 0 $\log_{10}$ copies/ml post-symptom onset. Unlike in experimental studies, this estimate for duration of shedding was not defined by assay detection limits. To estimate the average DFSO on which SARS-CoV-2 concentration reached diagnostic levels, we extrapolated the model pre-symptom onset to the equivalent of 1 and 3 $\log_{10}$ copies/ml (chosen as example assay detection limits) in specimen concentration for NPSs immersed in 1 ml of transport media, as described by the dilution factor estimation above. The average time from respiratory infection to reach diagnostic levels was then calculated by subtracting these values from the estimated average incubation period. Notably, the extrapolated time for SARS-CoV-2 to reach diagnostic concentrations in the respiratory tract should be validated in tracing studies, in which contacts are prospectively subjected to daily sampling.

**Calculation of rate profiles of expelled particles.** Rate profiles (particles/min or particles/cough) were calculated based on the corrected normalized concentration ($dC_n/d\log D_p$, in units of particles/cm$^3$) at each discrete particle size, normalization (32 size channels per decade) for the aerodynamic particle sizer used, unit conversion (cm$^3$ to L) and the sample flow rate (1 L/min). For coughing, the calculation assumed that participants coughed 10 times in the 30-s sampling interval. To determine the corrected normalized concentrations for breathing, we used a particle dilution factor of 4 and evaporation diameter factor of 0.5 (55). Breathing was taken to expel negligible quantities of larger respiratory particles based on the bronchiolar fluid film burst mechanism (55). To account for intermittent breathing while talking and singing, the rate profiles for these activities included the contribution of aerosols expelled by breathing.

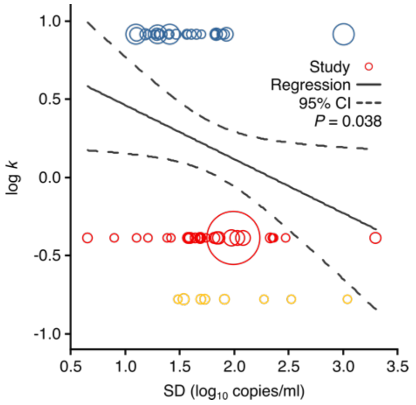


**Fig. S1.** Meta-regression between dispersion in SARS-CoV-2, SARS-CoV-1 and A(H1N1)pdm09 transmissibility and heterogeneity in rVL. Meta-regression of dispersion parameter ($k$) with the standard deviation (SD) of respiratory viral loads (rVLs) from all contributing studies (Pearson's $r$ = -0.26). Pooled estimates of $k$ were determined from the literature. Blue, red and yellow circles denote A(H1N1)pdm09 ($N$ = 27), COVID-19 ($N$ = 29) and SARS ($N$ = 8) studies, respectively. Circle sizes denote weighting in the meta-regression. The $P$-value was obtained using the meta-regression slope $t$-test.

| Subgroup, study ref. | Cases No. | Cases Type | Specimens No. | Specimens DFSO | Specimens Type | Respiratory viral load, $\log_{10}$ copies/ml **(95% CI)** | ***P* value for difference** |
|---|---|---|---|---|---|---|---|
| SARS-CoV-2 | | | | | | | |
| 62 | 4 | H, A, S | 9 | 4 to 10 | NPS, OPS | 4.12 (3.18-5.05) | |
| 42 | 102 | H, C, A, P, S, Ps, As | 109 | -2 to 10 | NPS, OPS | 4.63 (4.34-4.92) | |
| 63 | 6 | H, A, S | 6 | 3 to 8 | OPS | 5.13 (4.83-5.42) | |
| 83 | 77 | H, S, Ps | 119 | -1 to 8 | OPS, Spu | 5.26 (4.63-5.90) | |
| 43 | 23 | H, A, S | 51 | 0 to 10 | ETA, POS | 5.29 (4.65-5.93) | |
| 64 | 14 | H, A, S, As | 55 | 0 to 10 | NPS, OPS | 5.30 (4.87-5.74) | |
| 65 | - | H, N, A, S | 31 | 2 to 10 | NPS, OPS, Spu | 5.41 (4.81-6.01) | |
| 79 | 17 | H, A, P, S | 17 | 0 to 10 | NPS | 5.53 (4.36-6.71) | |
| 81 | 133 | C, A, P, S, Ps, As | 133 | -3 to 10 | NPS | 5.59 (5.31-5.88) | |
| 66 | - | H, A, S | 19 | 1 to 10 | POS, Spu | 5.62 (5.22-6.03) | |
| 80 | 7 | H, P, S, Ps, As | 14 | -1 to 10 | NPS | 5.69 (5.26-6.12) | |
| 67 | 2,200 | C, A, P, As | 2,200 | 0 | NPS | 6.01 (5.93-6.09) | |
| 68 | 171 | N, A, S | 171 | 0 to 10 | NPS | 6.07 (5.79-6.34) | |
| 69 | 103 | H, N, A, S | 103 | 0 to 8 | NPS, OPS | 6.10 (5.98-6.23) | |
| 21 | 9 | H, A, S, As | 133 | 2 to 10 | NPS, OPS, Spu | 6.21 (5.90-6.52) | |
| 70 | 41 | H, A, S | 310 | 2 to 10 | NPS, OPS, POS | 6.28 (6.05-6.50) | |
| 71 | 5 | H, A, S | 5 | 7 to 9 | NPS | 6.38 (5.16-7.60) | |
| 72 | - | H, A, S, As | 38 | 0 to 9 | NPA, NPS, OPS, Spu | 6.52 (5.77-7.27) | |
| 85 | 16 | H, A, P, S, Ps, As | 17 | -1 to 10 | NPS | 6.64 (5.83-7.45) | |
| 73 | 40 | H, A, P, S, As | 42 | 0 to 10 | NPS | 6.67 (6.17-7.18) | |
| 74 | 405 | H, A, P, S | 405 | 0 to 5 | NPS | 6.95 (6.78-7.17) | |
| 53 | 23 | H, P, S | 23 | 0 to 4 | NPS | 7.20 (6.23-8.16) | |
| 75 | 24 | H, A, S | 33 | 0 to 10 | NPS | 7.51 (6.97-8.05) | |
| 76 | 205 | H, N, A, S | 205 | 0 to 10 | NPS | 7.55 (7.30-7.81) | |
| 38 | - | H, A, S | 154 | 0 to 10 | NPS, Spu | 7.83 (7.58-8.08) | |
| 82 | 2 | H, A, P, S | 8 | 4 to 10 | NPS, OPS, Spu | 7.91 (6.69-9.12) | |
| 77 | 148 | N, A, S | 296 | 1 to 8 | NPS | 8.04 (7.81-8.28) | |
| 84 | 12 | H, P, S, As | 27 | 0 to 10 | NPS | 8.07 (7.47-8.67) | |
| 78 | 5 | H, A, S | 63 | 0 to 10 | NPS, OPS | 8.34 (7.95-8.73) | |
| Overall ($Q_{28} > 100$, $P < 0.001$, $I^2 = 97.1\%$) | | | | | | 6.34 (6.02-6.67) | - |
| SARS-CoV-1 | | | | | | | |
| 86 | 40 | H, A, S | 40 | 1 to 3 | NPA | 4.18 (3.24-5.12) | |
| 87 | 153 | H, A, S | 153 | 0 to 19 | NPS | 4.90 (4.64-5.16) | |
| 88 | 11 | H, A, S | 11 | 20 | NPS | 5.62 (4.12-7.11) | |
| 44 | - | H, A, S | 43 | 1 to 10 | NPA | 5.62 (4.94-6.30) | |
| 89 | 57 | H, A, S | 57 | 0 to 6 | NPA | 5.92 (5.43-6.42) | |
| 90 | 60 | H, A, S | 60 | 10 to 15 | NPA | 6.65 (6.21-7.08) | |
| 91 | 59 | H, A, S | 59 | 10 | NPA | 6.67 (6.23-7.11) | |
| 92 | 14 | H, A, S | 42 | 5 to 15 | NPA | 6.73 (6.28-7.18) | |
| Overall ($Q_7 > 100$, $P < 0.001$, $I^2 = 93.7\%$) | | | | | | 5.83 (5.11-6.55) | 0.111 |
| Influenza A(H1N1)pdm09 | | | | | | | |
| 93 | 97 | C, A, P, S, Ps, As | 218 | -2 to 9 | NPS | 5.28 (5.14-5.49) | |
| 94 | 51 | C, A, P, S, As | 129 | 1 to 7 | NPA, NPS, OPS | 5.31 (4.99-5.62) | |
| 95 | 581 | H, A, P, S | 581 | 1 to 8 | OPS | 5.46 (5.38-5.55) | |
| 96 | 13 | H, A, P, S | 25 | 2 to 9 | NPS | 5.69 (4.97-6.41) | |
| 97 | 17 | C, A, P, S | 20 | 0 to 6 | NPS, OPS | 5.73 (5.17-6.29) | |
| 98 | 27 | H, P, S | 41 | 2 to 7 | NPS | 5.84 (5.41-6.27) | |
| 99 | 64 | H, N, A, P, S | 89 | 1 to 7 | NPS | 6.18 (5.79-6.58) | |
| 100 | 533 | H, A, S | 533 | 0 to 9 | NPS | 6.29 (6.18-6.40) | |
| 101 | 633 | H, N, A, P, S | 533 | 0 to 3 | OPS | 6.31 (6.08-6.55) | |
| 102 | 47 | H, P, S | 47 | 1 to 4 | NPA | 6.49 (6.01-6.98) | |
| 103 | 12 | H, C, A, P, S | 21 | 3 to 5 | NPS | 6.83 (6.16-7.51) | |
| 104 | 48 | H, A, S | 48 | 0 to 7 | NPA | 6.84 (6.44-7.23) | |
| 105 | 58 | H, A, S | 58 | 0 to 9 | NPA, NPS, OPS | 6.86 (6.52-7.20) | |
| 106 | 87 | H, N, A, P, S | 87 | 0 to 7 | NPA, NPS | 6.87 (6.60-7.15) | |
| 107 | 33 | C, A, P, S, Ps, As | 123 | -1 to 8 | NPS, OPS | 7.16 (6.95-7.37) | |
| 108 | 34 | H, A, P, S | 34 | 0 to 2 | NPS | 7.18 (6.77-7.59) | |
| 109 | 74 | H, P, S | 282 | 1 to 9 | NPS | 7.22 (6.99-7.44) | |
| 110 | 578 | H, A, P, S | 578 | 1 to 7 | NPS | 7.22 (7.11-7.34) | |
| 111 | 27 | H, A, P, S | 59 | 0 to 7 | NPA, NPS, OPS | 7.24 (6.87-7.62) | |
| 112 | 45 | N, C, A, P, S | 54 | 0 to 8 | NPS, OPS | 7.28 (6.85-7.71) | |
| 113 | 22 | H, A, P, S | 22 | 0 to 7 | NPA, NPS, OPS | 7.29 (6.53-8.05) | |
| 114 | 86 | N, C, A, P, S, As | 86 | 0 to 8 | NPA, NPS, OPS | 7.32 (6.94-7.71) | |
| 115 | 60 | H, A, P, S | 60 | 0 to 3 | NPA | 7.45 (7.05-7.85) | |
| 116 | 12 | H, A, P, S | 33 | 2 to 9 | NPA, NPS, OPS | 7.59 (7.01-8.17) | |
| 117 | 50 | H, A, S | 50 | 0 to 9 | NPA, NPS | 7.94 (7.41-8.47) | |
| 118 | 209 | H, C, P, S | 209 | 0 to 7 | NPS | 8.21 (8.06-8.36) | |
| 119 | 251 | H, P, S | 251 | 0 to 2 | NPA | 8.82 (8.68-8.95) | |
| Overall ($Q_{26} > 100$, $P < 0.001$, $I^2 = 98.9\%$) | | | | | | 6.81 (6.41-7.22) | 0.040 |

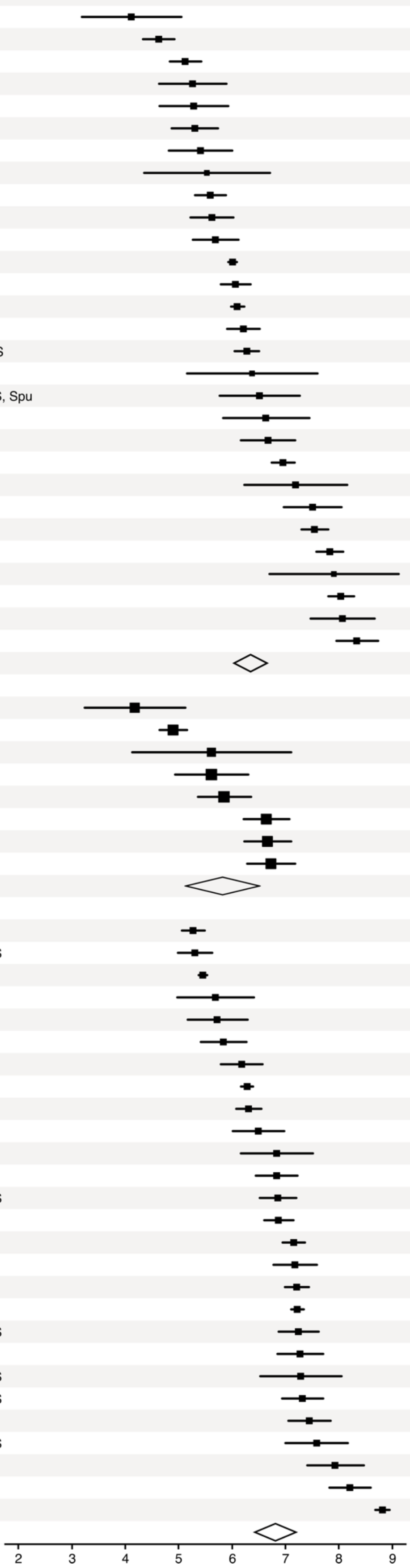

**Fig. S2.** Meta-analysis of rVLs of SARS-CoV-2, SARS-CoV-1 and influenza A(H1N1)pdm09 during the infectious period. Random-effects meta-analyses comparing the expected respiratory viral loads (rVLs) for COVID-19, SARS and A(H1N1)pdm09 cases during the infectious period. Quantitative specimen measurements were used to estimate rVLs, which refer to virus concentrations in the respiratory tract. Case types: hospitalized (H), not admitted (N), community (C), adult (A), pediatric (P), symptomatic (S), presymptomatic (Ps) and asymptomatic (As). Specimen types: endotracheal aspirate (ETA), nasopharyngeal aspirate (NPA), nasopharyngeal swab (NPS), oropharyngeal swab (OPS), posterior oropharyngeal saliva (POS) and sputum (Spu). Dashes denote case numbers that were not obtained. Box sizes denote weighting in the overall estimates. Between-study heterogeneity was assessed using the $P$-value from Cochran's $Q$ test and the $I^2$ statistic. References before 58 are listed in the main body. One-sided Welch's $t$-tests compared the expected SARS-CoV-2 rVL with those of SARS-CoV-1 and A(H1N1)pdm09 (non-significance, $P > 0.05$).

| Subgroup, study ref. | Cases No. | Cases Type | Specimens No. | Specimens DFSO | Specimens Type | Respiratory viral load, $\log_{10}$ copies/ml (95% CI) | P value for difference |
|---|---|---|---|---|---|---|---|
| SARS-CoV-2, low risk of bias | | | | | | | |
| 62 | 4 | H, A, S | 9 | 4 to 10 | NPS, OPS | 4.12 (3.18-5.05) | |
| 42 | 102 | H, C, A, P, S, Ps, As | 109 | -2 to 10 | NPS, OPS | 4.63 (4.34-4.92) | |
| 63 | 6 | H, A, S | 6 | 3 to 8 | OPS | 5.13 (4.83-5.42) | |
| 43 | 23 | H, A, S | 51 | 0 to 10 | ETA, POS | 5.29 (4.65-5.93) | |
| 64 | 14 | H, A, S, As | 55 | 0 to 10 | NPS, OPS | 5.30 (4.87-5.74) | |
| 65 | - | H, N, A, S | 31 | 2 to 10 | NPS, OPS, Spu | 5.41 (4.81-6.01) | |
| 79 | 17 | H, A, P, S | 17 | 0 to 10 | NPS | 5.53 (4.36-6.71) | |
| 81 | 133 | C, A, P, S, Ps, As | 133 | -3 to 10 | NPS | 5.59 (5.31-5.88) | |
| 66 | - | H, A, S | 19 | 1 to 10 | POS, Spu | 5.62 (5.22-6.03) | |
| 80 | 7 | H, P, S, Ps, As | 14 | -1 to 10 | NPS | 5.69 (5.26-6.12) | |
| 67 | 2,200 | C, A, P, As | 2,200 | 0 | NPS | 6.01 (5.93-6.09) | |
| 68 | 171 | N, A, S | 171 | 0 to 10 | NPS | 6.07 (5.79-6.34) | |
| 21 | 9 | H, A, S, As | 133 | 2 to 10 | NPS, OPS, Spu | 6.21 (5.90-6.52) | |
| 70 | 41 | H, A, S | 310 | 2 to 10 | NPS, OPS, POS | 6.28 (6.05-6.50) | |
| 73 | 40 | H, A, P, S, As | 42 | 0 to 10 | NPS | 6.67 (6.17-7.18) | |
| 74 | 405 | H, A, P, S | 405 | 0 to 5 | NPS | 6.95 (6.78-7.17) | |
| 63 | 23 | H, P, S | 23 | 0 to 4 | NPS | 7.20 (6.23-8.16) | |
| 75 | 24 | H, A, S | 33 | 0 to 10 | NPS | 7.51 (6.97-8.05) | |
| 76 | 205 | H, N, A, S | 205 | 0 to 10 | NPS | 7.55 (7.30-7.81) | |
| 38 | - | H, A, S | 154 | 0 to 10 | NPS, Spu | 7.83 (7.58-8.08) | |
| 84 | 2 | H, A, P, S | 8 | 4 to 10 | NPS, OPS, Spu | 7.91 (6.69-9.12) | |
| 77 | 148 | N, A, S | 296 | 1 to 8 | NPS | 8.04 (7.81-8.28) | |
| 82 | 12 | H, P, S, As | 27 | 0 to 10 | NPS | 8.07 (7.47-8.67) | |
| 78 | 5 | H, A, S | 63 | 0 to 10 | NPS, OPS | 8.34 (7.95-8.73) | |
| Overall ($Q_{23} > 100$, $P < 0.001$, $I^2 = 97.5\%$) | | | | | | 6.38 (6.98-6.78) | - |
| SARS-CoV-2, high risk of bias | | | | | | | |
| 83 | 77 | H, S, Ps | 119 | -1 to 8 | OPS, Spu | 5.26 (4.63-5.90) | |
| 69 | 103 | H, N, A, S | 103 | 0 to 8 | NPS, OPS | 6.10 (5.98-6.23) | |
| 71 | 5 | H, A, S | 5 | 7 to 9 | NPS | 6.38 (5.16-7.60) | |
| 72 | - | H, A, S, As | 38 | 0 to 9 | NPA, NPS, OPS, Spu | 6.52 (5.77-7.27) | |
| 85 | 16 | H, A, P, S, Ps, As | 17 | -1 to 10 | NPS | 6.64 (5.83-7.45) | |
| Overall ($Q_4 = 11.0$, $P = 0.027$, $I^2 = 63.6\%$) | | | | | | 6.11 (5.68-6.55) | 0.199 |

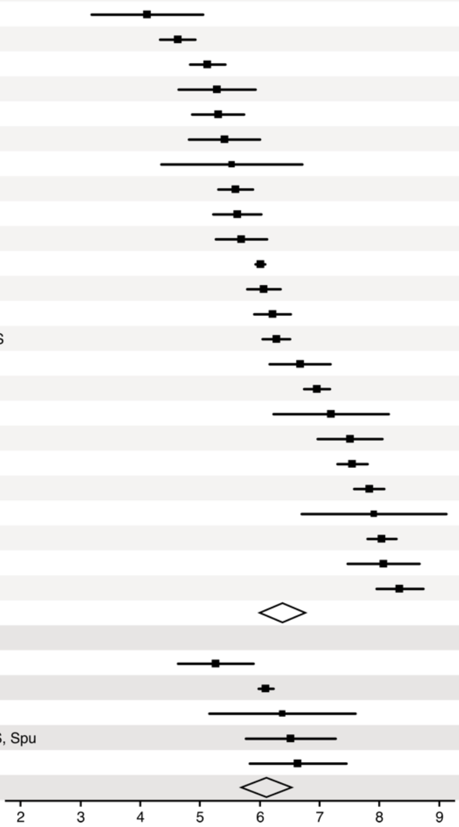


**Fig. S3.** Risk-of-bias sensitivity analysis of between-study heterogeneity for SARS-CoV-2 rVL during the infectious period. Random-effects meta-analyses, based on the risk of bias of contributing studies, of the expected respiratory viral loads (rVLs) of COVID-19 cases during the infectious period. Quantitative rVLs refer to virus concentrations in the respiratory tract. Case types: hospitalized (H), not admitted (N), community (C), adult (A), pediatric (P), symptomatic (S), presymptomatic (Ps) and asymptomatic (As). Specimen types: endotracheal aspirate (ETA), nasopharyngeal aspirate (NPA), nasopharyngeal swab (NPS), oropharyngeal swab (OPS), posterior oropharyngeal saliva (POS) and sputum (Spu). Dashes denote case numbers that were not obtained. Box sizes denote weighting in the overall estimates. One-sided Welch's *t*-test for difference (non-significance, $P > 0.05$). Between-study heterogeneity was assessed using the $P$-value from Cochran's $Q$ test (non-significance, $P > 0.05$) and the $I^2$ statistic ($I^2 < 30\%$ indicates low between-study heterogeneity). References before 58 are listed in the main body.

| Subgroup, study ref. | Cases No. | Cases Type | Specimens No. | Specimens DFSO | Specimens Type | Respiratory viral load, $log_{10}$ copies/ml (95% CI) | P value for difference |
|---|---|---|---|---|---|---|---|
| SARS-CoV-1, low risk of bias | | | | | | | |
| 87 | 153 | H, A, S | 153 | 0 to 19 | NPS | 4.90 (4.64-5.16) | |
| 88 | 11 | H, A, S | 11 | 20 | NPS | 5.62 (4.12-7.11) | |
| 44 | - | H, A, S | 43 | 1 to 10 | NPA | 5.62 (4.94-6.30) | |
| 89 | 57 | H, A, S | 57 | 0 to 6 | NPA | 5.92 (5.43-6.42) | |
| 90 | 60 | H, A, S | 60 | 10 to 15 | NPA | 6.65 (6.21-7.08) | |
| 91 | 59 | H, A, S | 59 | 10 | NPA | 6.67 (6.23-7.11) | |
| 92 | 14 | H, A, S | 42 | 5 to 15 | NPA | 6.73 (6.28-7.18) | |
| Overall ($Q_6$ = >100, $P < 0.001$, $I^2$ = 94.1%) | | | | | | 6.04 (5.31-6.76) | - |
| SARS-CoV-1, high risk of bias | | | | | | | |
| 86 | 40 | H, A, S | 40 | 1 to 3 | NPA | 4.18 (3.24-5.12) | - |

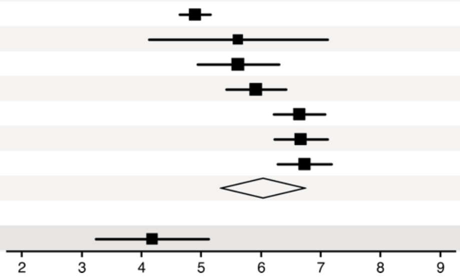


**Fig. S4.** Risk-of-bias sensitivity analysis of between-study heterogeneity for SARS-CoV-1 rVL during the infectious period. Random-effects meta-analyses, based on the risk of bias of contributing studies, of the expected respiratory viral loads (rVLs) of SARS cases during the infectious period. Quantitative rVLs refer to virus concentrations in the respiratory tract. Case types: hospitalized (H), adult (A) and symptomatic (S). Specimen types: nasopharyngeal aspirate (NPA) and nasopharyngeal swab (NPS). Dashes denote case numbers that were not obtained. Box sizes denote weighting in the overall estimates. One-sided Welch's *t*-test for difference, non-significance ($P > 0.05$). Between-study heterogeneity was assessed using the $P$-value from Cochran's $Q$ test (non-significance, $P > 0.05$) and the $I^2$ statistic ($I^2 < 30\%$ indicates low between-study heterogeneity). References before 58 are listed in the main body.

| Subgroup, study ref. | Cases No. | Cases Type | Specimens No. | Specimens DFSO | Specimens Type | Respiratory viral load, $log_{10}$ copies/ml (95% CI) | P value for difference |
|---|---|---|---|---|---|---|---|
| Influenza A(H1N1)pdm09, low risk of bias | | | | | | | |
| 93 | 97 | C, A, P, S, Ps, As | 218 | -2 to 9 | NPS | 5.28 (5.14-5.49) | |
| 94 | 51 | C, A, P, S, As | 129 | 1 to 7 | NPA, NPS, OPS | 5.31 (4.99-5.62) | |
| 95 | 581 | H, A, P, S | 581 | 1 to 8 | OPS | 5.46 (5.38-5.55) | |
| 96 | 13 | H, A, P, S | 25 | 2 to 9 | NPS | 5.69 (4.97-6.41) | |
| 97 | 17 | C, A, P, S | 20 | 0 to 6 | NPS, OPS | 5.73 (5.17-6.29) | |
| 98 | 27 | H, P, S | 41 | 2 to 7 | NPS | 5.84 (5.41-6.27) | |
| 99 | 64 | H, N, A, P, S | 89 | 1 to 7 | NPS | 6.18 (5.79-6.58) | |
| 100 | 533 | H, A, S | 533 | 0 to 9 | NPS | 6.29 (6.18-6.40) | |
| 102 | 47 | H, P, S | 47 | 1 to 4 | NPA | 6.49 (6.01-6.98) | |
| 103 | 12 | H, C, A, P, S | 21 | 3 to 5 | NPS | 6.83 (6.16-7.51) | |
| 104 | 48 | H, A, S | 48 | 0 to 7 | NPA | 6.84 (6.44-7.23) | |
| 105 | 58 | H, A, S | 58 | 0 to 9 | NPA, NPS, OPS | 6.86 (6.52-7.20) | |
| 106 | 87 | H, N, A, P, S | 87 | 0 to 7 | NPA, NPS | 6.87 (6.60-7.15) | |
| 107 | 33 | C, A, P, S, Ps, As | 123 | -1 to 8 | NPS, OPS | 7.16 (6.95-7.37) | |
| 109 | 74 | H, P, S | 282 | 1 to 9 | NPS | 7.22 (6.99-7.44) | |
| 110 | 578 | H, A, P, S | 578 | 1 to 7 | NPS | 7.22 (7.11-7.34) | |
| 111 | 27 | H, A, P, S | 59 | 0 to 7 | NPA, NPS, OPS | 7.24 (6.87-7.62) | |
| 112 | 45 | N, C, A, P, S | 54 | 0 to 8 | NPS, OPS | 7.28 (6.85-7.71) | |
| 115 | 60 | H, A, P, S | 60 | 0 to 3 | NPA | 7.45 (7.05-7.85) | |
| 116 | 12 | H, A, P, S | 33 | 2 to 9 | NPA, NPS, OPS | 7.59 (7.01-8.17) | |
| 117 | 50 | H, A, S | 50 | 0 to 9 | NPA, NPS | 7.94 (7.41-8.47) | |
| 119 | 251 | H, P, S | 251 | 0 to 2 | NPA | 8.82 (8.68-8.95) | |
| Overall ($Q_{21} > 100$, $P < 0.001$, $I^2 = 99.0\%$) | | | | | | 6.71 (6.25-7.17) | - |
| Influenza A(H1N1)pdm09, high risk of bias | | | | | | | |
| 101 | 633 | H, N, A, P, S | 533 | 0 to 3 | OPS | 6.31 (6.08-6.55) | |
| 108 | 34 | H, A, P, S | 34 | 0 to 2 | NPS | 7.18 (6.77-7.59) | |
| 113 | 22 | H, A, P, S | 22 | 0 to 7 | NPA, NPS, OPS | 7.29 (6.53-8.05) | |
| 114 | 86 | N, C, A, P, S, As | 86 | 0 to 8 | NPA, NPS, OPS | 7.32 (6.94-7.71) | |
| 118 | 209 | H, C, P, S | 209 | 0 to 7 | NPS | 8.21 (8.06-8.36) | |
| Overall ($Q_4 > 100$, $P < 0.001$, $I^2 = 96.5\%$) | | | | | | 7.27 (6.45-8.08) | 0.143 |

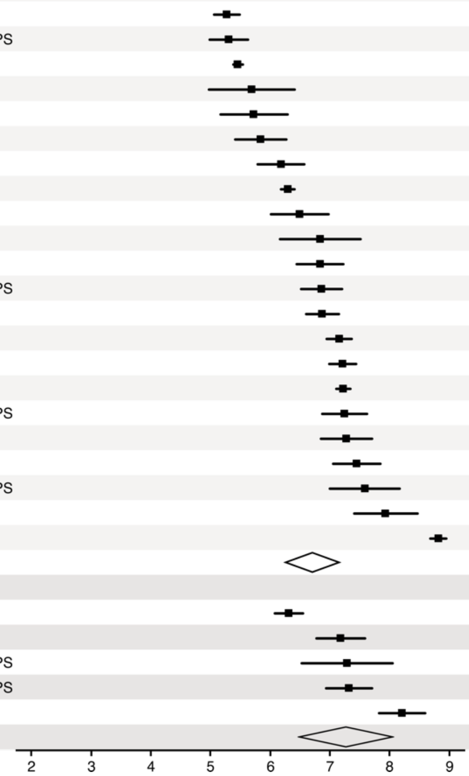


**Fig. S5.** Risk-of-bias sensitivity analysis of between-study heterogeneity for A(H1N1)pdm09 rVL during the infectious period. Random-effects meta-analyses, based on the risk of bias of contributing studies, of the expected respiratory viral loads (rVLs) of A(H1N1)pdm09 cases during the infectious period. Quantitative rVLs refer to virus concentrations in the respiratory tract. Case types: hospitalized (H), not admitted (N), community (C), adult (A), pediatric (P), symptomatic (S), presymptomatic (Ps) and asymptomatic (As). Specimen types: nasopharyngeal aspirate (NPA), nasopharyngeal swab (NPS) and oropharyngeal swab (OPS). Dashes denote case numbers that were not obtained. Box sizes denote weighting in the overall estimates. References before 58 are listed in the main body. One-sided Welch's *t*-test for difference (non-significance, $P > 0.05$). Between-study heterogeneity was assessed using the *P*-value from Cochran's *Q* test (non-significance, $P > 0.05$) and the $I^2$ statistic ($I^2 < 30\%$ indicates low between-study heterogeneity).

| Subgroup, study ref. | Cases No. | Cases Type | Specimens No. | Specimens DFSO | Specimens Type | Respiratory viral load, $\log_{10}$ copies/ml (95% CI) | P value for difference |
|---|---|---|---|---|---|---|---|
| Adult COVID-19, low risk of bias | | | | | | | |
| 62 | 4 | H, A, S | 9 | 4 to 10 | NPS, OPS | 4.12 (3.18-5.05) | |
| 42 | 100 | H, C, A, S, Ps, As | 106 | -2 to 10 | NPS, OPS | 4.65 (4.35-4.95) | |
| 63 | 6 | H, A, S | 6 | 3 to 8 | OPS | 5.13 (4.83-5.42) | |
| 43 | 23 | H, A, S | 51 | 0 to 10 | ETA, POS | 5.29 (4.65-5.93) | |
| 64 | 14 | H, A, S, As | 55 | 0 to 10 | NPS, OPS | 5.30 (4.87-5.74) | |
| 65 | - | H, N, A, S | 31 | 2 to 10 | NPS, OPS, Spu | 5.41 (4.81-6.01) | |
| 66 | - | H, A, S | 19 | 1 to 10 | POS, Spu | 5.62 (5.22-6.03) | |
| 67 | 2,182 | C, A, As | 2,182 | 0 | NPS | 6.01 (5.93-6.09) | |
| 68 | 171 | N, A, S | 171 | 0 to 10 | NPS | 6.07 (5.79-6.34) | |
| 21 | 9 | H, A, S, As | 133 | 2 to 10 | NPS, OPS, Spu | 6.21 (5.90-6.52) | |
| 70 | 41 | H, A, S | 310 | 2 to 10 | NPS, OPS, POS | 6.28 (6.05-6.50) | |
| 73 | 39 | H, A, S, As | 41 | 0 to 10 | NPS | 6.71 (6.20-7.23) | |
| 74 | 347 | H, A, S | 347 | 0 to 5 | NPS | 6.97 (6.77-7.18) | |
| 75 | 24 | H, A, S | 33 | 0 to 10 | NPS | 7.51 (6.97-8.05) | |
| 76 | 205 | H, N, A, S | 205 | 0 to 10 | NPS | 7.55 (7.30-7.81) | |
| 38 | - | H, A, S | 154 | 0 to 10 | NPS, Spu | 7.83 (7.58-8.08) | |
| 77 | 148 | N, A, S | 296 | 1 to 8 | NPS | 8.04 (7.81-8.28) | |
| 78 | 5 | H, A, S | 63 | 0 to 10 | NPS, OPS | 8.34 (7.95-8.73) | |
| Overall ($Q_{17} > 100$, $P < 0.001$, $I^2 = 98.0\%$) | | | | | | 6.32 (5.86-6.78) | - |
| Adult COVID-19, high risk of bias | | | | | | | |
| 69 | 103 | H, N, A, S | 103 | 0 to 8 | NPS, OPS | 6.10 (5.98-6.23) | |
| 71 | 5 | H, A, S | 5 | 7 to 9 | NPS | 6.38 (5.16-7.60) | |
| 72 | - | H, A, S, As | 38 | 0 to 9 | NPA, NPS, OPS, Spu | 6.52 (5.77-7.27) | |
| Overall ($Q_2 = 2.00$, $P = 0.502$, $I^2 = 0.0\%$) | | | | | | 6.11 (6.00-6.23) | 0.203 |

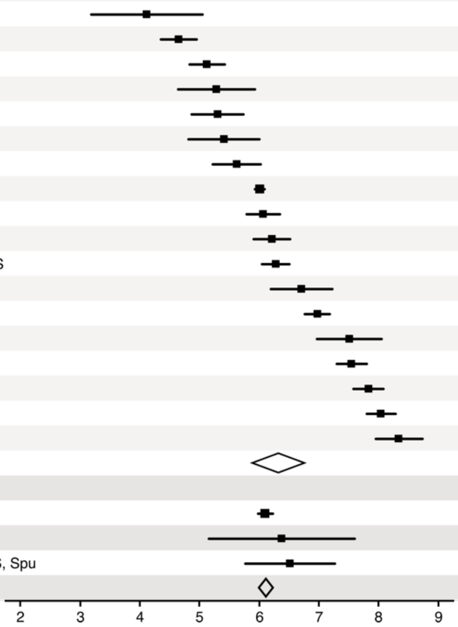


**Fig. S6.** Risk-of-bias sensitivity analysis of between-study heterogeneity for SARS-CoV-2 rVL for adult COVID-19 cases during the infectious period. Random-effects meta-analyses, based on the risk of bias of contributing studies, of the expected respiratory viral loads (rVLs) of adult (≥18 years old) COVID-19 cases during the infectious period. Quantitative rVLs refer to virus concentrations in the respiratory tract. Case types: hospitalized (H), not admitted (N), community (C), adult (A), pediatric (P), symptomatic (S), presymptomatic (Ps) and asymptomatic (As). Specimen types: endotracheal aspirate (ETA), nasopharyngeal aspirate (NPA), nasopharyngeal swab (NPS), oropharyngeal swab (OPS), posterior oropharyngeal saliva (POS) and sputum (Spu). Dashes denote case numbers that were not obtained. Box sizes denote weighting in the overall estimates. References before 58 are listed in the main body. One-sided Welch's *t*-test for difference (non-significance, $P > 0.05$). Between-study heterogeneity was assessed using the *P*-value from Cochran's *Q* test (non-significance, $P > 0.05$) and the $I^2$ statistic ($I^2 < 30\%$ indicates low between-study heterogeneity).

| Subgroup, study ref. | Cases No. | Cases Type | Specimens No. | Specimens DFSO | Specimens Type | Respiratory viral load, $log_{10}$ copies/ml (95% CI) | P value for difference |
|---|---|---|---|---|---|---|---|
| Symptomatic/presymptomatic COVID-19, low risk of bias | | | | | | | |
| 62 | 4 | H, A, S | 9 | 4 to 10 | NPS, OPS | 4.12 (3.18-5.05) | |
| 42 | 53 | H, C, A, P, S, Ps | 60 | -2 to 10 | NPS, OPS | 4.43 (4.03-4.84) | |
| 63 | 6 | H, A, S | 6 | 3 to 8 | OPS | 5.13 (4.83-5.42) | |
| 64 | 13 | H, A, S | 51 | 0 to 10 | NPS, OPS | 5.25 (4.79-5.71) | |
| 43 | 23 | H, A, S | 51 | 0 to 10 | ETA, POS | 5.29 (4.65-5.93) | |
| 65 | - | H, N, A, S | 31 | 2 to 10 | NPS, OPS, Spu | 5.41 (4.81-6.01) | |
| 79 | 17 | H, A, P, S | 17 | 0 to 10 | NPS | 5.53 (4.36-6.71) | |
| 66 | - | H, A, S | 19 | 1 to 10 | POS, Spu | 5.62 (5.22-6.03) | |
| 80 | 6 | H, P, S, Ps | 13 | -1 to 10 | NPS | 5.77 (5.10-6.53) | |
| 81 | 81 | C, A, P, S, Ps | 81 | -3 to 10 | NPS | 5.78 (5.41-6.15) | |
| 21 | 8 | H, A, S | 129 | 2 to 10 | NPS, OPS, Spu | 5.88 (5.60-6.17) | |
| 68 | 171 | N, A, S | 171 | 0 to 10 | NPS | 6.07 (5.79-6.34) | |
| 70 | 41 | H, A, S | 310 | 2 to 10 | NPS, OPS, POS | 6.28 (6.05-6.50) | |
| 73 | 31 | H, A, P, S | 33 | 2 to 10 | NPS | 6.95 (6.37-7.53) | |
| 74 | 405 | H, A, P, S | 405 | 0 to 5 | NPS | 6.95 (6.78-7.17) | |
| 53 | 23 | H, P, S | 23 | 0 to 4 | NPS | 7.20 (6.23-8.16) | |
| 75 | 24 | H, A, S | 33 | 0 to 10 | NPS | 7.51 (6.97-8.05) | |
| 76 | 205 | H, N, A, S | 205 | 0 to 10 | NPS | 7.55 (7.30-7.81) | |
| 38 | - | H, A, S | 154 | 0 to 10 | NPS, Spu | 7.83 (7.58-8.08) | |
| 84 | 2 | H, A, P, S | 8 | 4 to 10 | NPS, OPS, Spu | 7.91 (6.69-9.12) | |
| 77 | 148 | N, A, S | 296 | 1 to 8 | NPS | 8.04 (7.81-8.28) | |
| 82 | 9 | H, P, S | 20 | 0 to 10 | NPS | 8.10 (7.37-8.33) | |
| 78 | 5 | H, A, S | 63 | 0 to 10 | NPS, OPS | 8.34 (7.95-8.73) | |
| Overall ($Q_{22}$ > 100, $P$ < 0.001, $I^2$ = 91.7%) | | | | | | 6.43 (5.92-6.93) | - |
| Symptomatic/presymptomatic COVID-19, high risk of bias | | | | | | | |
| 83 | 77 | H, S, Ps | 119 | -1 to 8 | OPS, Spu | 5.25 (4.79-5.71) | |
| 69 | 103 | H, N, A, S | 103 | 0 to 8 | NPS, OPS | 6.10 (5.98-6.23) | |
| 71 | 5 | H, A, S | 5 | 7 to 9 | NPS | 6.38 (5.16-7.60) | |
| Overall ($Q_2$ = 2.0, $P$ = 0.421, $I^2$ = 0.0%) | | | | | | 6.14 (5.90-6.38) | 0.167 |

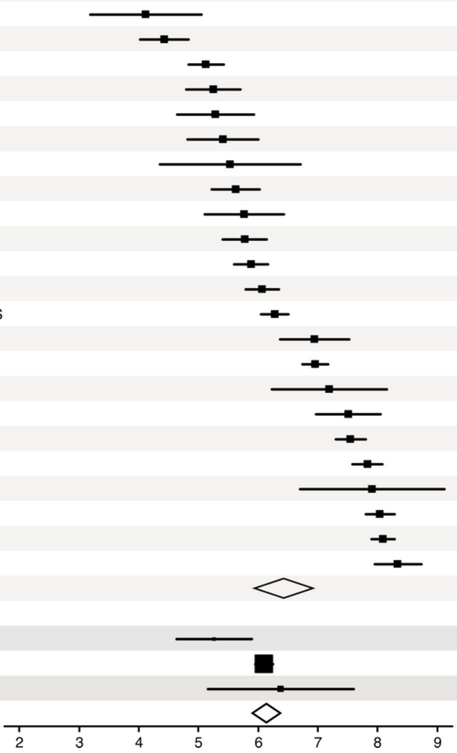


**Fig. S7.** Risk-of-bias sensitivity analysis of between-study heterogeneity for SARS-CoV-2 rVL for symptomatic/presymptomatic COVID-19 cases during the infectious period. Random-effects meta-analyses, based on the risk of bias of contributing studies, of the expected respiratory viral loads (rVLs) of symptomatic/presymptomatic (≥18 years old) COVID-19 cases during the infectious period. Quantitative rVLs refer to virus concentrations in the respiratory tract. Case types: hospitalized (H), not admitted (N), community (C), adult (A), pediatric (P), symptomatic (S), presymptomatic (Ps) and asymptomatic (As). Specimen types: endotracheal aspirate (ETA), nasopharyngeal swab (NPS), oropharyngeal swab (OPS), posterior oropharyngeal saliva (POS) and sputum (Spu). Dashes denote case numbers that were not obtained. Box sizes denote weighting in the overall estimates. References before 58 are listed in the main body. One-sided Welch's *t*-test for difference (non-significance, $P$ > 0.05). Between-study heterogeneity was assessed using the $P$-value from Cochran's $Q$ test (non-significance, $P > 0.05$) and the $I^2$ statistic the $I^2$ statistic ($I^2 < 30\%$ indicates low between-study heterogeneity).

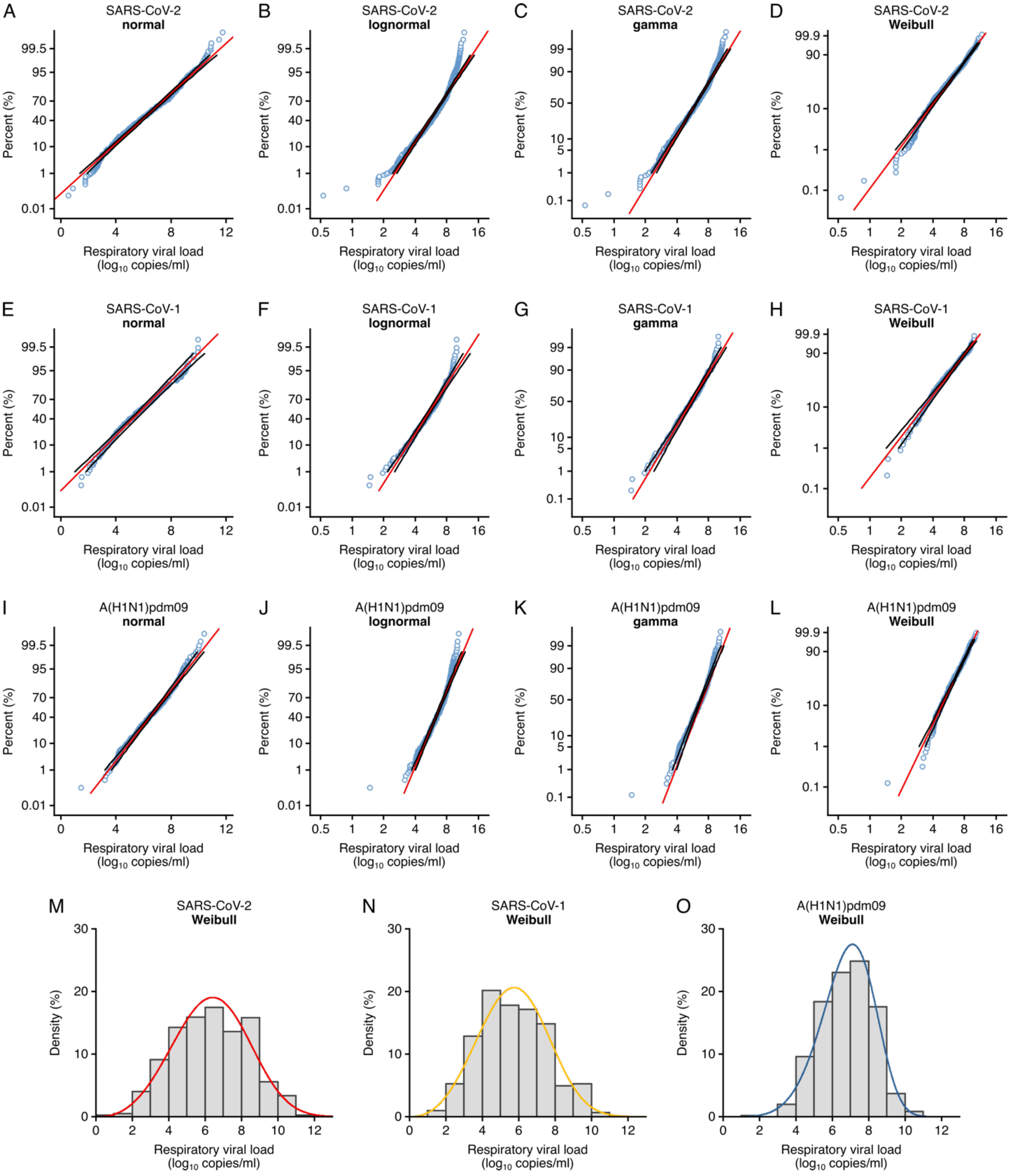


**Fig. S8.** Respiratory viral loads for SARS-CoV-2, SARS-CoV-1 and A(H1N1)pdm09 best conform to Weibull distributions. (*A* to *D*) Normal ($P \leq 0.01$) (*A*), lognormal ($P \leq 0.01$) (*B*), gamma ($P \leq 0.005$) (*C*) and Weibull ($P > 0.10$, not significant [NS]) (*D*) probability plots for individual sample data of SARS-CoV-2 rVLs across days from symptom onset in the systematic dataset ($N = 941$ samples from $N = 20$ studies). (*E* to *H*) Normal ($P > 0.05$, NS) (*E*), lognormal ($P \leq 0.01$) (*F*), gamma ($P > 0.05$, NS) (*G*) and Weibull ($P > 0.10$, NS) (*H*) probability plots for individual sample data of SARS-CoV-1 rVLs in the systematic dataset ($N = 303$ samples from $N = 5$ studies). (*I* to *L*) Normal ($P \leq 0.01$) (*I*), lognormal ($P \leq 0.01$) (*J*), gamma ($P \leq 0.005$) (*K*) and Weibull ($P > 0.10$, NS) (*L*) probability plots for individual sample data of A(H1N1)pdm09 rVLs in the systematic dataset ($N = 512$ samples from $N = 10$ studies). These categories included only rVL data from positive (above the detection limit) qRT-PCR measurements. The *P*-values were determined using the modified Kolmogorov-Smirnov test for the goodness of fit of each distribution. When the null hypothesis is accepted (NS at $P > 0.05$), the probability density function cannot be rejected to describe the distribution of the data. Blue circles, black lines and red lines represent individual sample data, expected distributions and 95% CIs, respectively. (*M* to *O*) Histograms and fitted Weibull distributions of the above data for SARS-CoV-2 (*M*), SARS-CoV-1 (*N*) and A(H1N1)pdm09 (*O*).

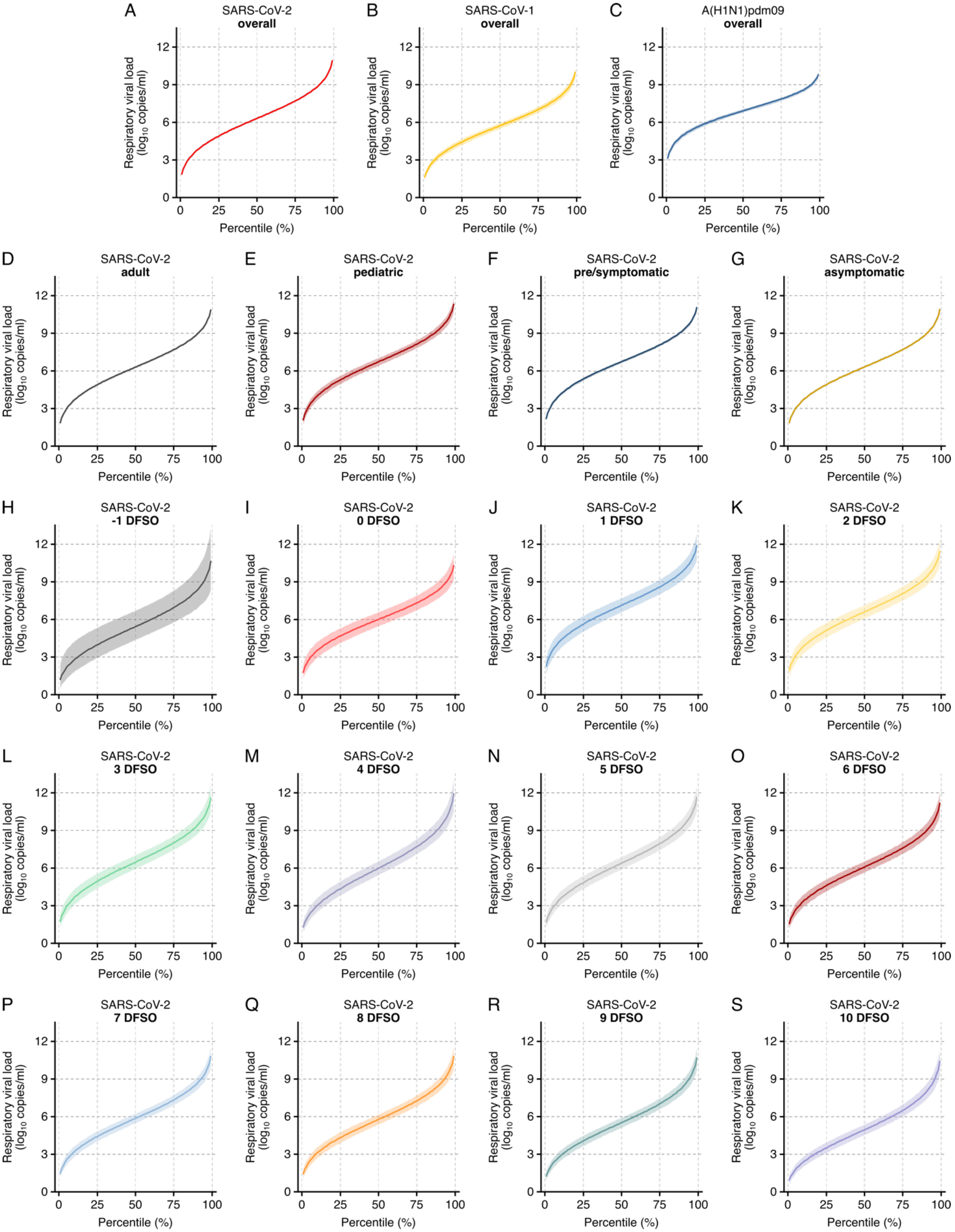
A SARS-CoV-2 overall
B SARS-CoV-1 overall
C A(H1N1)pdm09 overall
D SARS-CoV-2 adult
E SARS-CoV-2 pediatric
F SARS-CoV-2 pre/symptomatic
G SARS-CoV-2 asymptomatic
H SARS-CoV-2 -1 DFSO
I SARS-CoV-2 0 DFSO
J SARS-CoV-2 1 DFSO
K SARS-CoV-2 2 DFSO
L SARS-CoV-2 3 DFSO
M SARS-CoV-2 4 DFSO
N SARS-CoV-2 5 DFSO
O SARS-CoV-2 6 DFSO
P SARS-CoV-2 7 DFSO
Q SARS-CoV-2 8 DFSO
R SARS-CoV-2 9 DFSO
S SARS-CoV-2 10 DFSO
Respiratory viral load ($log_{10}$ copies/ml)
Percentile (%)

**Fig. S9.** Case heterogeneity in rVLs across viruses, COVID-19 subgroups and disease course. (*A* to *C*) Estimated respiratory viral loads (rVLs) of SARS-CoV-2 (*A*), SARS-CoV-1 (*B*) and A(H1N1)pdm09 (*C*) across case percentile (cp) throughout the infectious periods. (*D* to *G*) Estimated SARS-CoV-2 rVLs for adult (*D*), pediatric (*E*), symptomatic/presymptomatic (*F*) and asymptomatic (*G*) cases across cp throughout the infectious period. (*H* to *S*) Estimated SARS-CoV-2 rVLs across cp on different days from symptom onset (DFSO) during the infectious period. Earlier DFSO were excluded based on limited data. Data ranged between the 1$^{st}$ and 99$^{th}$ cps. Sample numbers, distribution parameters and descriptive statistics are summarized in *SI Appendix*, Table S7. Lines and bands represent estimates and 95% CIs, respectively.

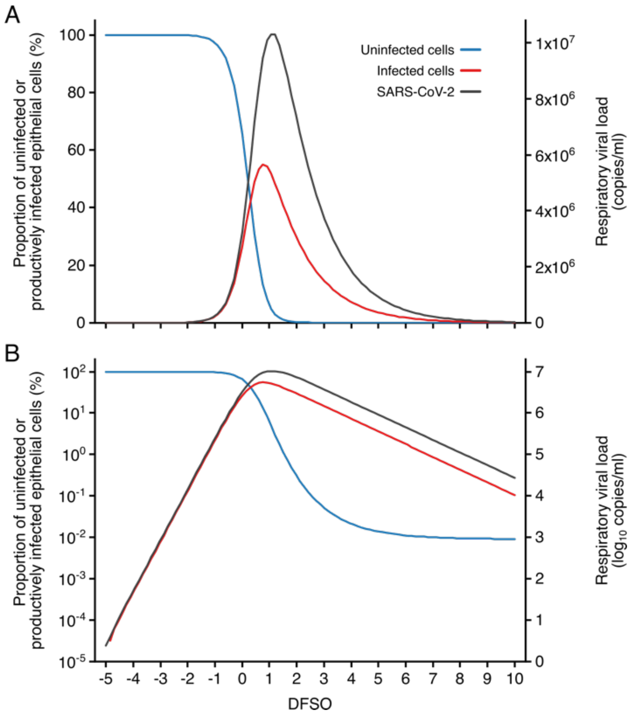


**Fig. S10.** Kinetics of SARS-CoV-2 and airway epithelial cells during respiratory infection. (*A* and *B*) Estimated kinetics of uninfected (blue) and productively infected (red) airway epithelial cells (left axis) and SARS-CoV-2 (right axis) in the respiratory tract, as shown in linear (*A*) and logarithmic (*B*) scales.

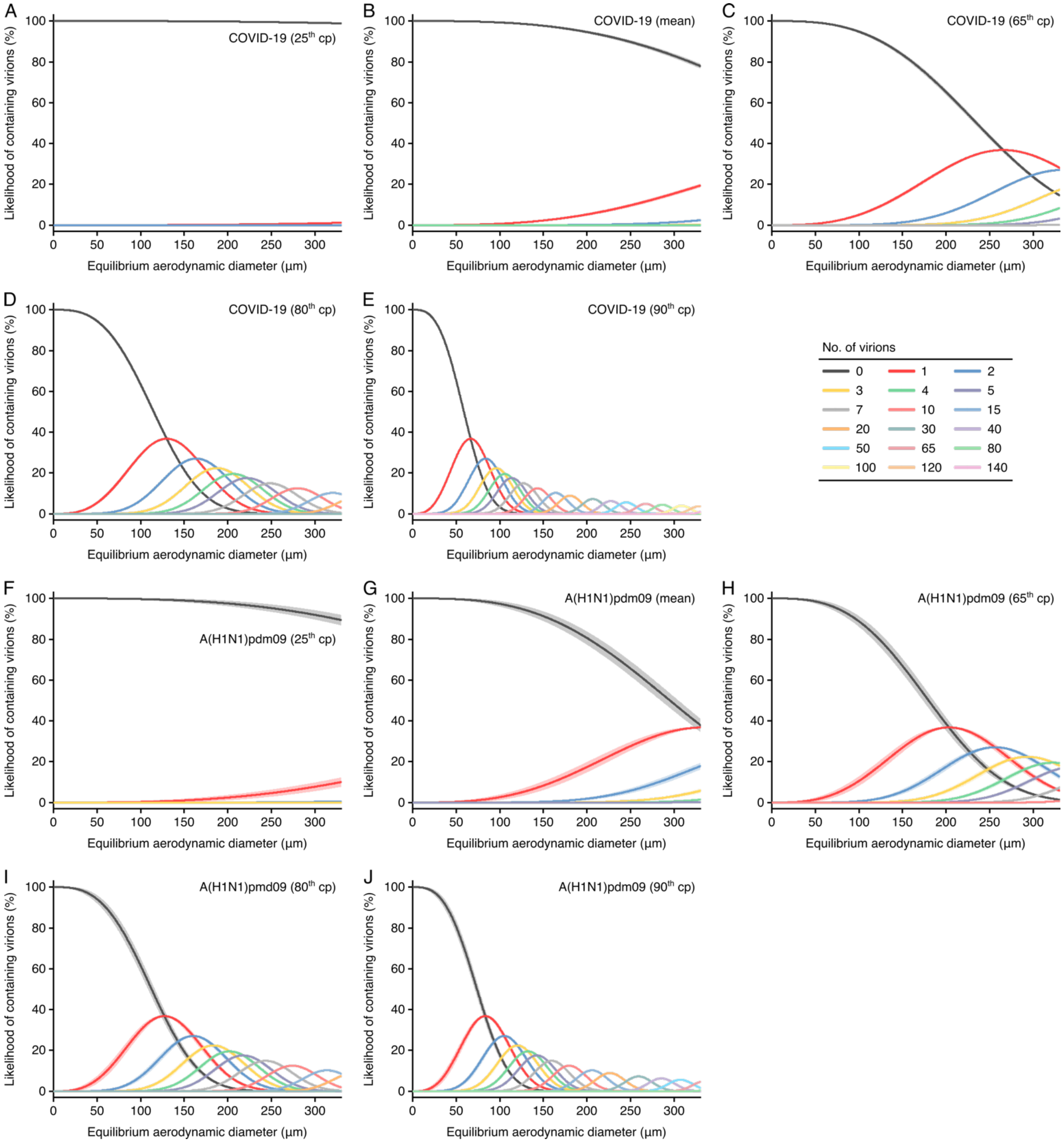


**Fig. S11.** Likelihood of respiratory particles containing SARS-CoV-2 or A(H1N1)pdm09. (*A* to *E*) Estimated likelihood that droplets and aerosols contain viable SARS-CoV-2 when expelled by the 25th case percentile (cp) (*A*), mean (*B*), 65th cp (*C*), 80th cp (*D*) or 90th cp (*E*) for COVID-19 cases during the infectious period. (*F* to *J*) Estimated likelihood that droplets and aerosols contain viable A(H1N1)pdm09 when expelled by the $25^{th}$ cp (*F*), mean (*G*), $65^{th}$ cp (*H*), $80^{th}$ cp (*I*) or $90^{th}$ cp (*J*) for A(H1N1)pmd09 cases during the infectious period. For higher no. of virions, some likelihood curves were omitted to aid visualization. When the likelihood for 0 virions approaches 0%, particles are expected to contain at least one viable copy. Lines and bands represent estimates and 95% CIs, respectively, for estimated likelihoods.

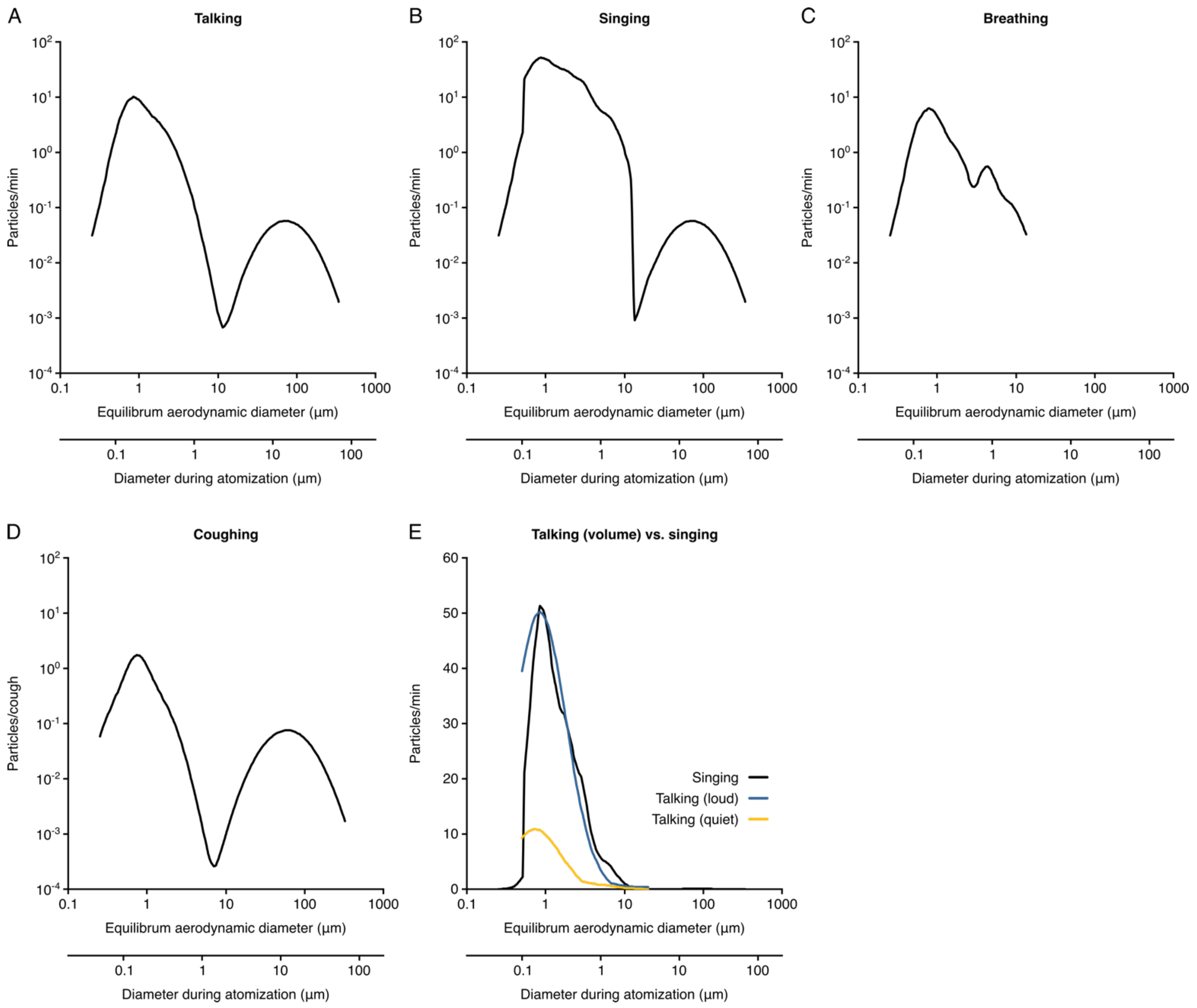


**Fig. S12.** Rate profiles for particle expelled by respiratory activities. (*A* to *D*) Rate profiles of particles expelled while talking (*A*), singing (*B*), breathing (*C*) and coughing (*D*). (*E*) Comparison of the rate profiles of aerosol emission from singing and different amplitudes of talking. The rate profles were calculated from the normalized concentrations in Johnson et al (55) (*A*, *B*, *D* and *E*) and Morawaska et al (56) (*C*) or collected from Asadi et al (57) (*E*). Dehydrated particle diameters were taken to be 0.5 times the hydrated diameter during atomization. Breathing was taken to expel negligible quantities of larger particles based on the bronchiolar fluid film burst mechanism.

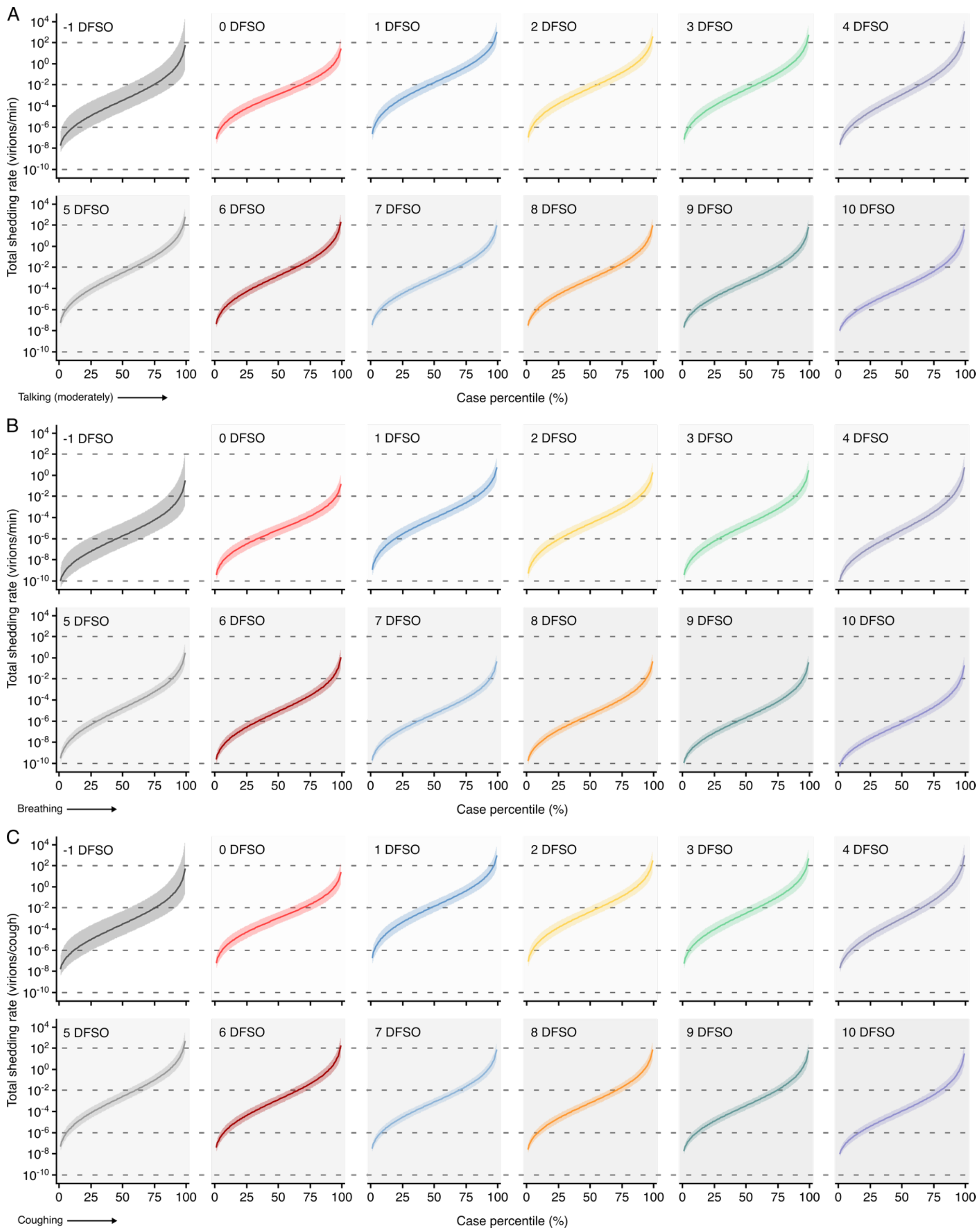


**Fig. S13.** Heterogeneity in shedding SAR-CoV-2 via talking, breathing and coughing. (*A* to *C*) Case heterogeneity in the total SARS-CoV-2 shedding rate (over all particle sizes) by talking at a moderate amplitude (*A*), breathing (*B*) or coughing (*C*) for COVID-19 cases across the infectious period. Earlier presymptomatic days were excluded based on limited data. Data represent estimated rates for viable virus and range between the 1$^{st}$ and 99$^{th}$ cps. Lines and bands represent estimates and 95% CIs, respectively.

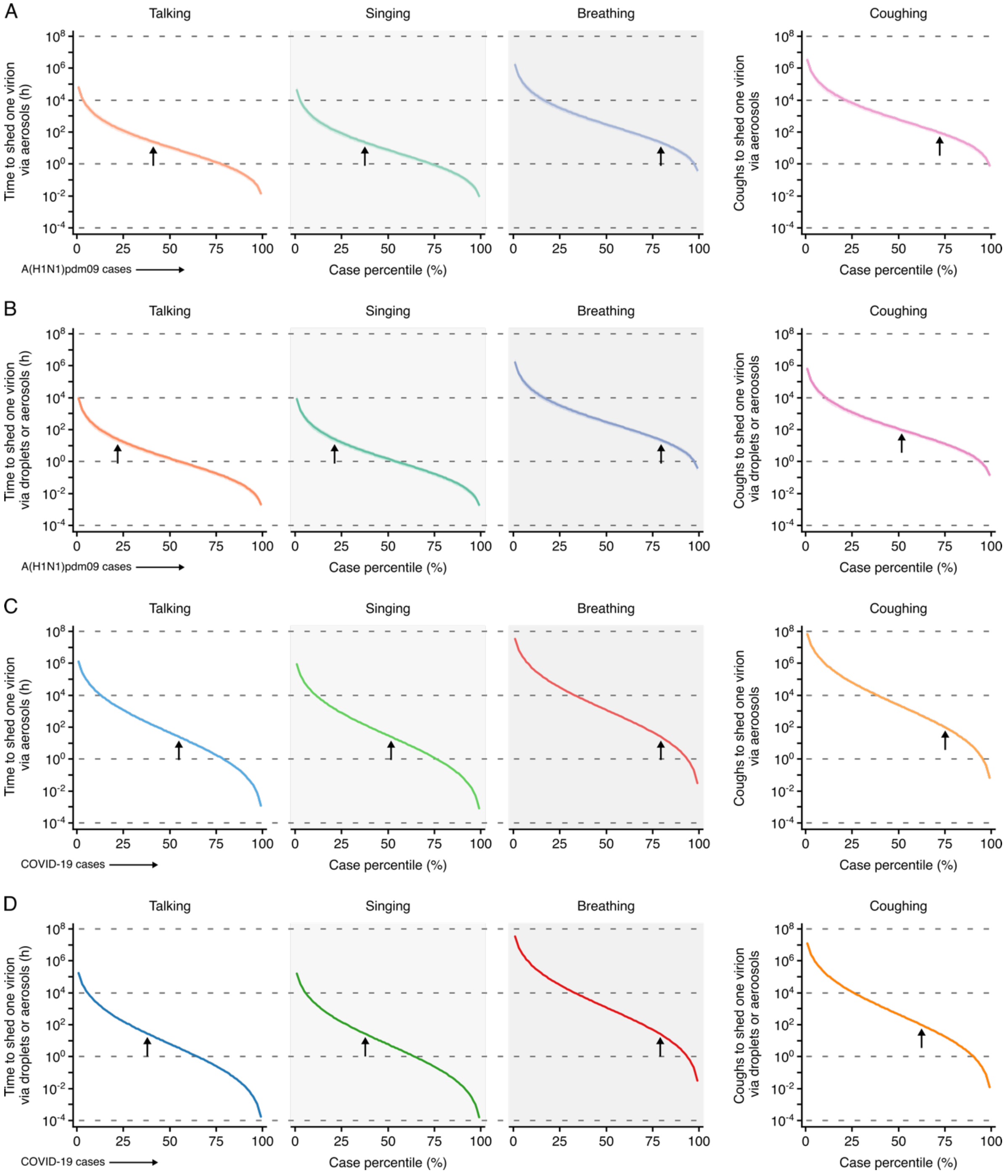


**Fig. S14.** Heterogeneity in infectiousness for COVID-19 and A(H1N1)pmd09 cases during the infectious period. (*A* and *B*) Estimated time for a A(H1N1)pdm09 case to expel one virion via only aerosols (*A*) or either droplets or aerosols (*B*) by talking, singing, breathing or coughing. (*C* and *D*) Estimated time for a COVID-19 case to expel one SARS-CoV-2 virion via only aerosols

(*C*) or either droplets or aerosols (*D*) by talking, singing, breathing or coughing. Data represent estimated times to expel viable virus and range between the $1^{st}$ and $99^{th}$ case percentiles (cps). Vertical arrows depict the cp expected to shed 1 virion in 24 h (talking, singing or breathing) or 100 coughs. Lines and bands represent estimates and 95% CIs, respectively.

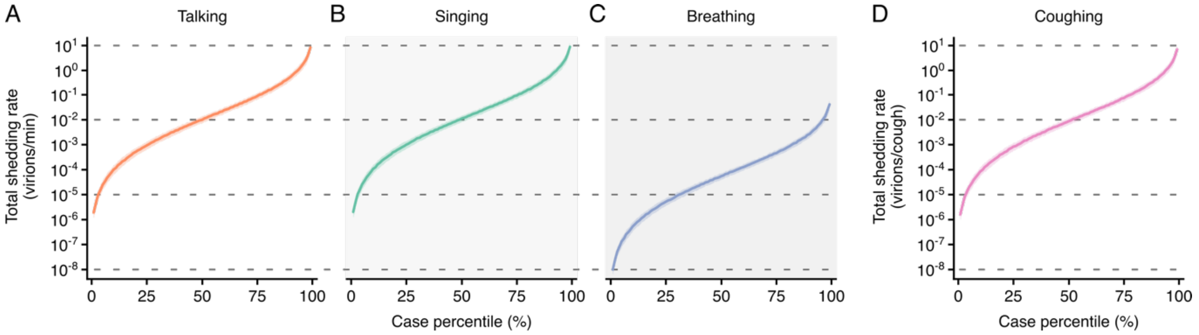


**Fig. S15.** Heterogeneity in shedding A(H1N1)pdm09 via droplets and aerosols. (*A* to *D*) Case heterogeneity in the total A(H1N1)pdm09 shedding rates while talking (*A*), singing (*B*), breathing (*C*) and coughing (*D*) during the infectious period. Data represent estimated rates for viable virus and range between the 1st and 99th cps. Lines and bands represent estimates and 95% CIs, respectively.

**Table S1.** Search strategy used for MEDLINE.

| Database: **Ovid MEDLINE® and Epub Ahead of Print, In-Process & Other Non-Indexed Citations and Daily** 1946 to August 07, 2020 | |
|---|---|
| **#** | **Searches** |
| 1 | exp Coronavirus/ |
| 2 | exp Coronavirus Infections/ |
| 3 | exp Betacoronavirus/ |
| 4 | (coronavirus* or corona virus* or betacoronavirus* or OC43 or NL63 or 229E or HKU1 or HcoV* or ncov* or covid* or sars-cov* or sarscov* or Sars-coronavirus* or Severe Acute Respiratory Syndrome* or sudden acute respiratory syndrome*).tw,kf. |
| 5 | (2019nCov* or 2019-novel CoV or corona or covid19 or ((novel or new or nouveau) adj2 (CoV or Pandemi*))).tw,kf. |
| 6 | (pneumonia.tw,kf. Or exp pneumonia/) and (Wuhan or Hubei).tw,kf. |
| 7 | COVID-19.rx,px,ox. Or severe acute respiratory syndrome coronavirus 2.os. |
| 8 | exp Influenza A Virus, H1N1 Subtype/ |
| 9 | (“A/H1N1*” or H1N1* or pdm09 or ((influenza or virus or pandemic) adj4 “2009”) or influenza A or swine flu).tw,kf. |
| 10 | 1 or 2 or 3 or 4 or 5 or 6 or 7 or 8 or 9 |
| 11 | ((respiratory adj3 (specimen* or sample* or swab*)) or sputum or nares or endotrachea* or endotrache* or endotra* or ((nasal or oral* or throat) adj3 (swab* or sample* or smear* or specimen*)) or NPS or OPS or ((endotrachea* or endotracheal*) adj2 aspirat*) or NPA or ETA or (deep adj4 saliva) or POS or “swab sample*” or “flocked swab*”).tw,kf. |
| 12 | Nasal cavity/vi |
| 13 | Sputum/vi |
| 14 | Nasopharynx/vi |
| 15 | Oropharynx/vi |
| 16 | *Saliva/vi |
| 17 | Pharynx/vi |
| 18 | (clinical adj2 (sample* or specimen*)).tw,kf. |
| 19 | (“RT-PCR” or “RTPCR” or “ddPCR” or “polymerase chain reaction”).tw,kf. |
| 20 | Influenza, Human/vi |
| 21 | exp Coronavirus Infections/vi or exp Coronavirus/vi or exp Betacoronavirus/vi |
| 22 | polymerase chain reaction/ or multiplex polymerase chain reaction/ or real-time polymerase chain reaction/ or reverse transcriptase polymerase chain reaction/ |
| 23 | 11 or 12 or 13 or 14 or 15 or 16 or 17 or 18 or 19 or 20 or 21 or 22 |
| 24 | 10 and 23 |
| 25 | (vir* load* or vir* shed* or vir* burden or vir* titer* or vir* titre* or (vir* adj2 count*)).tw,kf. |
| 26 | (((copies or copy) adj2 (ml* or milli* or microl*)) or ((RNA* or vir*) adj2 concentration*)).tw,kf. |

| | |
|---|---|
| 27 | ((calibration adj1 curve*) or (standard adj1 curve*)).tw,kf. |
| 28 | ((ct* adj1 value*) or cycle threshold or (copies adj2 test*) or (copy adj2 test*) or ((copy or copies) adj2 number*)).tw,kf. |
| 29 | Viral load/ |
| 30 | Viral shedding/ |
| 31 | (“copy/m*” or “copies/m*” or “copy/test*” or “copies/test*”).tw,kf. |
| 32 | ((test or diagnos*) adj2 sensitiv*).tw,kf. |
| 33 | 25 or 26 or 27 or 28 or 29 or 30 or 31 or 32 |
| 34 | 24 and 33 |
| 35 | animals/ not humans/ |
| 36 | 34 not 35 |

**Table S2.** Search strategy used for EMBASE.

| Database: **Embase** 1974 to 2020 August 06 | |
|---|---|
| **#** | **Searches** |
| 1 | coronavirus infection/ or severe acute respiratory syndrome/ |
| 2 | COVID 19/ |
| 3 | coronavirus disease 2019/ |
| 4 | severe acute respiratory syndrome coronavirus 2/ or SARS coronavirus 2/ |
| 5 | SARS coronavirus/ or betacoronavirus/ |
| 6 | “influenza a virus (h1n1)”/ |
| 7 | 2009 h1n1 influenza/ or “influenza a (h1n1)”/ |
| 8 | (coronavirus* or corona virus* or betacoronavirus* or OC43 or NL63 or 229E or HKU1 or HcoV* or ncov* or covid* or sars-cov* or sarscov* or Sars-coronavirus* or Severe Acute Respiratory Syndrome* or Sudden acute respiratory syndrome*).tw,kw. |
| 9 | (2019nCov* or 2019-novel CoV or corona or covid19 or ((novel or new or nouveau) adj2 (CoV or Pandemi*))).tw,kw. |
| 10 | (pneumonia.tw,kw. Or exp pneumonia/) and (Wuhan or Hubei).tw,kw. |
| 11 | (covid or SARS or H1N1 or coronavirus).ox. |
| 12 | (“A/H1N1*” or H1N1* or pdm09 or ((influenza or virus or pandemic) adj4 “2009”) or influenza A or swine flu).tw,kw. |
| 13 | 1 or 2 or 3 or 4 or 5 or 6 or 7 or 8 or 9 or 10 or 11 or 12 |
| 14 | nose smear/ |
| 15 | smear/ or nose smear/ or sputum smear/ |
| 16 | *sputum/ |
| 17 | *nasopharynx/ or *pharynx/ |
| 18 | throat culture/ |
| 19 | saliva analysis/ |
| 20 | *oropharynx/ |
| 21 | exp nasopharyngeal aspiration/ |
| 22 | real time polymerase chain reaction/ or real time reverse transcription polymerase chain reaction/ or reverse transcription polymerase chain reaction/ |
| 23 | (clinical adj2 (sample* or specimen*)).tw,kw. |
| 24 | ((respiratory adj3 (specimen* or sample* or swab*)) or sputum or nares or endotrachea* or endotrache* or endotra* or ((nasal or oral* or throat) adj3 (swab* or sample* or smear* or specimen*)) or NPS or OPS or ((endotrachea* or endotracheal*) adj2 aspirat*) or NPA or ETA or (deep adj4 saliva) or POS or “swab sample*” or “flocked swab*”).tw,kw. |
| 25 | (“RT-PCR” or “RTPCR” or “ddPCR” or “polymerase chain reaction”).tw,kw. |
| 26 | *2009 H1N1 influenza/di or coronavirus disease 2019/di or Coronavirus infection/di |

| | |
|---|---|
| 27 | 14 or 15 or 16 or 17 or 18 or 19 or 20 or 21 or 22 or 23 or 24 or 25 or 26 |
| 28 | 13 and 27 |
| 29 | virus load/ |
| 30 | virus shedding/ |
| 31 | (vir* load* or vir* shed* or vir* burden or vir* titer* or vir* titre* or (vir* adj2 count*)).tw,kw. |
| 32 | (((copies or copy) adj2 (ml* or milli* or microl*)) or ((RNA* or vir*) adj2 concentration*)).tw,kw. |
| 33 | ((calibration adj1 curve*) or (standard adj1 curve*)).tw,kw. |
| 34 | ((ct* adj1 value*) or cycle threshold or (copies adj2 test*) or (copy adj2 test*) or ((copy or copies) adj2 number*)).tw,kw. |
| 35 | (“copy/m*” or “copies/m*” or “copy/test*” or “copies/test*”).tw,kw. |
| 36 | ((test or diagnos*) adj2 sensitiv*).tw,kw. |
| 37 | 29 or 30 or 31 or 32 or 33 or 34 or 35 or 36 |
| 38 | 28 and 37 |
| 39 | exp animal/ not human/ |
| 40 | 38 not 39 |

**Table S3.** Search strategy used for Cochrane Central.

| Database(s): **EBM Reviews – Cochrane Central Register of Controlled Trials** July 2020 | |
|---|---|
| **#** | **Searches** |
| 1 | exp Coronavirus/ |
| 2 | exp Coronavirus Infections/ |
| 3 | betacoronavirus/ |
| 4 | (coronavirus* or corona virus* or betacoronavirus* or OC43 or NL63 or 229E or HKU1 or HcoV* or ncov* or covid* or sars-cov* or sarscov* or Sars-coronavirus* or Severe Acute Respiratory Syndrome* or sudden acute respiratory syndrome*).tw,kw. |
| 5 | (2019nCov* or 2019-novel CoV or corona or covid19 or ((novel or new or nouveau) adj2 (CoV or Pandemi*))).tw,kw. |
| 6 | (pneumonia.tw,kw. Or exp pneumonia/) and (Wuhan or Hubei).tw,kw. |
| 7 | exp Influenza A Virus, H1N1 Subtype/ |
| 8 | ("A/H1N1*" or H1N1* or pdm09 or ((influenza or virus or pandemic) adj4 "2009") or influenza A or swine flu).tw,kw. |
| 9 | 1 or 2 or 3 or 4 or 5 or 6 or 7 or 8 |
| 10 | ((respiratory adj3 (specimen* or sample* or swab*)) or sputum or nares or endotrachea* or endotrache* or endotra* or ((nasal or oral* or throat) adj3 (swab* or sample* or smear* or specimen*)) or NPS or OPS or ((endotrachea* or endotracheal*) adj2 aspirat*) or NPA or ETA or (deep adj4 saliva) or POS or "swab sample*" or "flocked swab*").tw,kw. |
| 11 | Nasal cavity/ |
| 12 | Sputum/ |
| 13 | Nasopharynx/ |
| 14 | Oropharynx/ |
| 15 | *Saliva/ |
| 16 | Pharynx/ |
| 17 | (clinical adj2 (sample* or specimen*)).tw,kw. |
| 18 | ("RT-PCR" or "RTPCR" or "ddPCR" or "polymerase chain reaction").tw,kw. |
| 19 | exp Coronavirus Infections/vi or exp Coronavirus/vi or exp Betacoronavirus/vi |
| 20 | polymerase chain reaction/ or multiplex polymerase chain reaction/ or real-time polymerase chain reaction/ or reverse transcriptase polymerase chain reaction/ |
| 21 | 10 or 11 or 12 or 13 or 14 or 15 or 16 or 17 or 18 or 19 or 20 |
| 22 | 9 and 21 |
| 23 | (vir* load* or vir* shed* or vir* burden or vir* titer* or vir* titre* or (vir* adj2 count*)).tw,kw. |
| 24 | (((copies or copy) adj2 (ml* or milli* or microl*)) or ((RNA* or vir*) adj2 concentration*)).tw,kw. |
| 25 | ((calibration adj1 curve*) or (standard adj1 curve*)).tw,kw. |

| | |
|---|---|
| 26 | ((ct* adj1 value*) or cycle threshold or (copies adj2 test*) or (copy adj2 test*) or ((copy or copies) adj2 number*)).tw,kw. |
| 27 | Viral load/ |
| 28 | Viral shedding/ |
| 29 | (“copy/m*” or “copies/m*” or “copy/test*” or “copies/test*”).tw,kw. |
| 30 | ((test or diagnos*) adj2 sensitiv*).tw,kw. |
| 31 | 23 or 24 or 25 or 26 or 27 or 28 or 29 or 30 |
| 32 | 22 and 31 |

**Table S4.** Search strategy used for Web of Science Core Collection.

| **Web of Science Core Collection** | |
|---|---|
| **#** | **Searches** |
| #1 | TOPIC: ((coronavirus* or “corona virus*” or betacoronavirus* or OC43 or NL63 or 229E or HKU1 or HcoV* or ncov* or covid* or “sars-cov*” or sarscov* or “Sars-coronavirus*” or “Severe Acute Respiratory Syndrome*” or “sudden acute respiratory syndrome*” or “2019-ncov*” or 2019nCov* or “2019-novel CoV” or corona or ((novel or new or nouveau) NEAR/2 (CoV or Pandemi*) ) OR (pneumonia and (Wuhan or Hubei) ) or “A/H1N1*” or H1N1* or pdm09 or ((influenza or virus or pandemic) NEAR/4 “2009”) or “influenza A” or “swine flu”))<br>Indexes=SCI-EXPANDED, SSCI, A&HCI, CPCI-S, CPCI-SSH, ESCI Timespan=All years |
| #2 | TOPIC: (((respiratory NEAR/3 (specimen* or sample* or swab*) ) or sputum or nares or endotrachea* or endotrache* or endotra* or ((nasal or oral* or throat) NEAR/3 (swab* or sample* or smear* or specimen*) ) or NPS or OPS or ((endotrachea* or endotracheal*) NEAR/2 aspirat*) or NPA or ETA or (deep NEAR/4 saliva) or POS or “swab sample*” or “flocked swab*” or (clinical NEAR/2 (sample* or specimen*) ) or “RT-PCR” or “RTPCR” or “ddPCR” or “polymerase chain reaction”))<br>Indexes=SCI-EXPANDED, SSCI, A&HCI, CPCI-S, CPCI-SSH, ESCI Timespan=All years |
| #3 | TOPIC: ((“vir* load*” or “vir* shed*” or “vir* burden” or “vir* titer*” or “vir* titre*” or (vir* NEAR/2 count*) or ((copies or copy) NEAR/2 (ml or mls or milli* or microl*) ) or ((RNA* or vir*) NEAR/2 concentration*) or (calibration NEAR/1 curve*) or (standard NEAR/1 curve*) or “ct value*” or “cycle threshold” or ((copies or copy) NEAR/2 test*) or ((copy or copies) NEAR/2 number*) or “copy/m*” or “copies/m*” or “copy/test*” or “copies/test*” or ((test or diagnos*) NEAR/2 sensitiv*)))<br>Indexes=SCI-EXPANDED, SSCI, A&HCI, CPCI-S, CPCI-SSH, ESCI Timespan=All years |
| #4 | #3 AND #2 AND #1<br>Refined by: [excluding] WEB OF SCIENCE CATEGORIES: ( VETERINARY SCIENCES )<br>Indexes=SCI-EXPANDED, SSCI, A&HCI, CPCI-S, CPCI-SSH, ESCI Timespan=All years |

**Table S5.** Search strategy used for medRxiv and bioRxiv.

| **medRxiv + bioRxiv (via Publish or Perish program)** |
|---|
| **Keywords:** (covid OR coronavirus OR ncov OR hcov OR h1n1 OR “swine flu” OR COVID19 or SARS) AND (“copies/ml” OR “copy/ml” OR “viral load” OR “copies/test” OR “copy/test” OR “copies per ml” OR “copy per ml”)<br>**Publication name:** MedRxiv |
| **Keywords:** (covid OR coronavirus OR ncov OR hcov OR h1n1 OR “swine flu” OR COVID19 or SARS) AND (“copies/ml” OR “copy/ml” OR “viral load” OR “copies/test” OR “copy/test” OR “copies per ml” OR “copy per ml”)<br>**Publication name:** BioRxiv |

**Table S6.** Characteristics of contributing studies.

| Study* | Country | No. of cases included (no. of specimens) | No. of pediatric cases (no. of specimens) | No. of asymptomatic cases (no. of specimens) | Disease caused by virus | Case definition (WHO) | Pharmaco-therapy (type)† | Individual data extracted (diluent volu-me reported)‡ | Adjusted viral load§ (type of specimen) | Weight, % (meta-analysis category)‖ | Weight, % (meta-regression) | Risk of bias¶ |
|---|---|---|---|---|---|---|---|---|---|---|---|---|
| Argyropoulos et al. (2020) (76) | USA | 205 (205) | 0 | 0 | COVID-19 | Confirmed | No | No (no) | Yes (NPS) | 3.80 (V), 5.09 (A), 3.98 (S/Ps) | 2.13 | ******** |
| Baggio et al. (2020) (74) | Switzerland | 405 (405) | 58 (58) | 0 | COVID-19 | Confirmed | No | Yes (no) | Yes (NPS) | 3.84 (V), 5.14 (A), 13.9 (P), 3.88 (S/Ps) | 4.20 | ******* |
| Fajnzylber et al. (2020) (65) | USA | - (31) | 0 | 0 | COVID-19 | Confirmed | No | Yes (yes) | Yes (NPS, OPS)<br>No (Spu) | 3.45 (V), 4.62 (A), 3.76 (S/Ps) | 0.32 | ******** |
| Han et al. (2020) (84) | South Korea | 2 (8) | 1 (6) | 0 | COVID-19 | Confirmed | No | Yes (no) | Yes (NPS, OPS) | 2.53 (V), 4.06 (S/Ps) | 0.08 | ****** |
| Han et al. (2020) (82) | South Korea | 12 (27) | 12 (27) | 3 (7) | COVID-19 | Confirmed | No | Yes (no) | Yes (NPS) | 3.43 (V), 17.5 (P), 4.10 (S/Ps), 14.3 (As) | 0.28 | ******** |
| Hung et al. (2020) (70) | China | 41 (310) | 0 | 0 | COVID-19 | Confirmed | No (control group) | No (no) | Yes (NPS, OPS, POS) | 3.81 (V), 5.10 (A), 3.81 (S/Ps) | 3.22 | ******** |
| Hurst et al. (2020) (81) | USA | 133 (133) | 54 (54) | 52 (52) | COVID-19 | Confirmed | No | Yes (no) | Yes (NPS) | 3.77 (V), 15.3 (P), 3.88 (S/Ps), 21.6 (As) | 1.38 | ******** |
| Iwasaki et al. (2020) (71) | Japan | 5 (5) | 0 | 0 | COVID-19 | Confirmed | No | Yes (no) | Yes (NPS) | 2.53 (V), 3.37 (A), 4.12 (S/Ps) | 0.05 | *** |
| Kawasuji et al. (2020) (85) | Japan | 16 (16) | - | - | COVID-19 | Confirmed | Yes (antivirals - type not reported) | Yes (no) | Yes (NPS) | 3.15 (V) | 0.18 | ***** |
| L'Huillier et al. (2020) (53) | Switzerland | 23 (23) | 23 (23) | 0 | COVID-19 | Confirmed | No | Yes (no) | Yes (NPS) | 2.91 (V), 14.7 (P), 3.73 (S/Ps) | 0.24 | ******** |
| Lavezzo et al. (2020) (42) | Italy | 103 (110) | 2 (3) | 49 (49) | COVID-19 | Confirmed | No | Yes (yes) | Yes (NPS, OPS) | 3.77 (V), 5.03 (A), 11.57 (P), 3.57 (S/Ps), 21.8 (As) | 1.14 | ******* |
| Lennon et al. (2020) (67) | USA | 2,200 (2,200) | 18 (18) | 2,200 (2,200#) | COVID-19 | Confirmed | No | No (yes) | Yes (NPS) | 3.88 (V), 5.20 (A), 24.0 (As) | 22.84 | ******** |
| Lucas et al. (2020) (75) | USA | 24 (33) | 0 | 0 | COVID-19 | Confirmed | Moderate and severe patients (tocilizumab) | Yes (yes) | Yes (NPS) | 3.51 (V), 4.69 (A), 4.08 (S/Ps) | 0.34 | ******* |
| Mitja et al. (2020) (77) | Spain | 148 (296) | 0 | 0 | COVID-19 | Confirmed | No (control group) | No (no) | Yes (NPS) | 3.81 (V), 5.10 (A), 3.93 (S/Ps) | 3.07 | ******** |

| | | | | | | | | | | | | |
|---|---|---|---|---|---|---|---|---|---|---|---|---|
| Pan et al. (2020) (83) | China | 75 (104) | - | 0 | COVID-19 | Confirmed | No | Yes (no) | Yes (OPS)<br>No (Spu) | 3.45 (V), 2.50 (S/Ps) | 1.24 | **** |
| Peng et al. (2020) (63) | China | 6 (6) | 0 | 0 | COVID-19 | Confirmed | Yes (arbidol, lopinavir, ritonavir) | Yes (no) | Yes (OPS) | 3.03 (V), 4.05 (A), 4.02 (S/Ps) | 0.06 | ******** |
| Perera et al. (2020) (72) | China | - (36) | 0 | - | COVID-19 | Confirmed | No | Yes (no) | Yes (NPA, NPS, OPS, Spu) | 3.23 (V), 4.32 (A) | 0.39 | **** |
| Shi et al. (2020) (69) | China | 103 (103) | 0 | 0 | COVID-19 | Confirmed | No | Yes (no) | Yes (NPS, OPS) | 3.87 (V), 5.18 (A), 4.34 (S/Ps) | 1.07 | ***** |
| Shrestha et al. (2020) (68) | USA | 171 (171) | 0 | 0 | COVID-19 | Confirmed | No | Yes (no) | Yes (NPS) | 3.79 (V), 5.07 (A), 3.86 (S/Ps) | 1.78 | ******* |
| To et al. (2020) (43) | China | 23 (51) | 0 | 0 | COVID-19 | Confirmed | No | Yes (yes) | Yes (ETA, POS) | 3.37 (V), 4.51 (A), 3.25 (S/Ps) | 0.53 | ********* |
| van Kampen et al. (2020) (38) | The Netherlands | - (154) | 0 | 0 | COVID-19 | Confirmed | No | Yes (yes) | Yes (NPS, Spu) | 3.80 (V), 5.09 (A), 4.10 (S/Ps) | 1.60 | ******* |
| Vetter et al. (2020) (78) | Switzerland | 5 (63) | 0 | 0 | COVID-19 | Confirmed | No | Yes (yes) | Yes (NPS, OPS) | 3.68 (V), 4.93 (A), 4.14 (S/Ps) | 0.65 | ********* |
| Wölfel et al. (2020) (21) | Germany | 9 (136) | 0 | 1 (4) | COVID-19 | Confirmed | No | Yes (yes) | Yes (NPS, OPS)<br>No (Spu) | 3.76 (V), 5.03 (A), 3.93 (S/Ps) | 1.38 | ******* |
| Wyllie et al. (73) | USA | 40 (42) | - | 9 (9) | COVID-19 | Confirmed | No | Yes (yes) | Yes (NPS) | 3.55 (V), 4.75 (A), 4.00 (S/Ps), 18.3 (As) | 0.44 | ******* |
| Xu et al. (2020) (80) | China | 7 (14) | 7 (14) | 1 (1) | COVID-19 | Confirmed | No | Yes (no) | Yes (NPS) | 3.40 (V). 17.3 (P), 4.1 (S/Ps) | 0.15 | ******** |
| Yonker et al. (2020) (79) | USA | 17 (17) | 14 (14) | 0 | COVID-19 | Confirmed | No | Yes (no) | Yes (NPS) | 2.58 (V), 9.79 (P), 3.28 (S/Ps) | 0.18 | ****** |
| Zhang et al. (2020) (62) | China | 9 (9) | 0 | 0 | COVID-19 | Confirmed | No | Yes (no) | Yes (NPS, OPS) | 2.97 (V), 3.97 (A), 3.68 (S/Ps) | 0.09 | ******** |
| Zheng et al. (2020) (66) | China | - (19) | 0 | 0 | COVID-19 | Confirmed | No | Yes (no) | Yes (POS, Spu) | 3.66 (V), 4.90 (A), 4.23 (S/Ps) | 0.20 | ******* |
| Zou et al. (2020) (64) | China | 14 (55) | 0 | 1 (4) | COVID-19 | Confirmed | No | Yes (no) | Yes (NPS, OPS) | 3.64 (V), 4.87 (A), 3.65 (S/Ps) | 0.57 | ******** |
| Chen et al. (2006) (87) | China | 154 (154[#]) | 0 | 0 | SARS | Confirmed | Yes (oseltamivir, broad-spectrum antibiotics, ribavirin) | Yes (no) | Yes (NPS) | 14.0 (V) | 1.59 | ******** |
| Chu et al. (2004) (88)[☆] | China | 11 (11) | 0 | 0 | SARS | Confirmed | No (control group) | Yes (yes) | Yes (NPS) | 8.6 (V) | 0.11 | ********* |
| Chu et al. (2005) (89) | China | 57 (57) | 0 | 0 | SARS | Confirmed | No | Yes (yes) | Yes (NPA) | 13.3 (V) | 0.59 | ********* |
| Cheng et al. (2004) (91) | China | 59 (59) | 0 | 0 | SARS | Confirmed | Yes (ribavirin, hydrocortisone, | Yes (yes) | Yes (NPA) | 13.4 (V) | 0.61 | ******** |

| | | | | | | | | | | | |
|---|---|---|---|---|---|---|---|---|---|---|---|
| | | | | | | | prednisolone, methylpredni-solone) | | | | |
| Hung et al. (2004) (90) | China | 60 (60) | 0 | 0 | SARS | Confirmed | Yes (ribavirin, hydrocortisone, prednisolone, methylpredni-solone) | No (yes) | Yes (NPA) | 13.5 (V) | 0.62 | ******* |
| Peiris et al. (2003) (92)[☆] | China | 14 (42) | 0 | 0 | SARS | Confirmed | Yes (ribavirin, hydrocortisone, prednisolone, methylpredni-solone) | Yes (no) | Yes (NPA) | 13.4 (V) | 0.44 | ******** |
| Poon et al. (2003) (86) | China | 40 (40) | 0 | 0 | SARS | Confirmed | No | No (yes) | Yes (NPA) | 11.3 (V) | 0.42 | ***** |
| Poon et al. (2004) (44) | China | - (43) | 0 | 0 | SARS | Confirmed | - | No (yes) | Yes (NPA) | 12.5 (V) | 0.45 | ****** |
| Alves et al. (2020) (114) | Brazil | 86 (86) | - | 15 (15) | Influenza A(H1N1)pdm09 | Confirmed | No | No (yes) | Yes (NPA, NPS, OPS) | 3.7 (V) | 0.89 | ***** |
| Chan et al. (2011) (105) | China | 58 (58) | 0 | 0 | Influenza A(H1N1)pdm09 | Confirmed | No (pretreatment) | Yes (no) | Yes (NPA, NPS, OPS) | 3.7 (V) | 0.60 | ****** |
| Cheng et al. (2010) (115) | China | 60 (60) | - | 0 | Influenza A(H1N1)pdm09 | Confirmed | No (pretreatment) | No (no) | Yes (NPA) | 3.7 (V) | 0.62 | ****** |
| Cowling et al. (2010) (112) | China | 45 (54) | 22 (31) | 0 | Influenza A(H1N1)pdm09 | Confirmed | Yes (22 cases on oseltamivir) | Yes (yes) | Yes (NPS, OPS) | 3.7 (V) | 0.56 | ******* |
| Duchamp et al. (2010) (118) | France | 209 (209) | 209 (209) | 0 | Influenza A(H1N1)pdm09 | Confirmed | Yes (oseltamivir, zanamivir) | No (yes) | Yes (NPS) | 3.8 (V) | 2.17 | ***** |
| Esposito et al. (2011) (109) | Italy | 74 (282) | 74 (282) | 0 | Influenza A(H1N1)pdm09 | Confirmed | No | Yes (yes) | Yes (NPS) | 3.8 (V) | 2.93 | ****** |
| Hung et al. (2010) (106) | China | 87 (87) | - | 0 | Influenza A(H1N1)pdm09 | Confirmed | Yes (oseltamivir) | Yes (no) | Yes (NPA, NPS) | 3.8 (V) | 0.90 | ****** |
| Ip et al. (2016) (97) | China | 17 (20) | 7 (-) | 0 | Influenza A(H1N1)pdm09 | Confirmed | No | Yes (no) | Yes (NPS, OPS) | 3.6 (V) | 0.21 | ****** |
| Ito et al. (2012) (108) | Japan | 34 (34) | - | 0 | Influenza A(H1N1)pdm09 | Confirmed | No (pretreatment) | Yes (yes) | Yes (NPS) | 3.7 (V) | 0.35 | ***** |
| Killingley et al. (2010) (103) | United Kingdom | 12 (21) | - | 0 | Influenza A(H1N1)pdm09 | Confirmed | Yes (oseltamivir) | Yes (yes) | Yes (NPS) | 3.5 (V) | 0.22 | ******** |
| Launes et al. (2012) (102) | Spain | 47 (47) | 47 (47) | 0 | Influenza A(H1N1)pdm09 | Confirmed | No (pretreatment) | No (no) | Yes (NPA) | 3.7 (V) | 0.49 | ******* |
| Lee et al. (2011) (104) | China | 48 (48) | 0 | 0 | Influenza A(H1N1)pdm09 | Confirmed | No (pretreatment) | No (no) | Yes (NPA) | 3.7 (V) | 0.50 | ****** |
| Lee et al. (2011) (110) | Singapore | 578 (578) | 231 (231) | 0 | Influenza A(H1N1)pdm09 | Confirmed | No (pretreatment) | No (no) | Yes (NPS) | 3.8 (V) | 6.00 | ********* |

| | | | | | | | | | | | | |
|---|---|---|---|---|---|---|---|---|---|---|---|---|
| Li et al. (2010) (95) | China | 581 (581) | 522 (522) | 0 | Influenza A(H1N1)pdm09 | Confirmed | No (pretreatment) | No (no) | Yes (OPS) | 3.8 (V) | 6.03 | ****** |
| Li et al. (2010) (111) | China | 27 (59) | - | 0 | Influenza A(H1N1)pdm09 | Confirmed | No (non-treated group) | No (no) | Yes (NPA, NPS, OPS) | 3.7 (V) | 0.61 | ******* |
| Loeb et al. (2012) (93) | Canada | 97 (218) | - | - (17) | Influenza A(H1N1)pdm09 | Confirmed | No | No (no) | Yes (NPS) | 3.8 (V) | 2.26 | ******* |
| Lu et al. (2012) (96) | China | 13 (25) | - | 0 | Influenza A(H1N1)pdm09 | Confirmed | Yes (oseltamivir, zanamivir) | Yes (no) | Yes (NPS) | 3.5 (V) | 0.26 | ******* |
| Meschi et al. (2011) (99) | Italy | 533 (533) | 0 | 0 | Influenza A(H1N1)pdm09 | Confirmed | No (pretreatment) | No (no) | Yes (NPS) | 3.8 (V) | 0.92 | ********* |
| Ngaosuwankul et al. (2010) (116) | China | 12 (33) | - | 0 | Influenza A(H1N1)pdm09 | Confirmed | No (pretreatment) | No (yes) | Yes (NPA, NPS, OPS) | 3.6 (V) | 0.34 | ****** |
| Rath et al. (2012) (98) | Germany | 27 (41) | 27 (41) | 0 | Influenza A(H1N1)pdm09 | Confirmed | Yes (oseltamivir) | Yes (yes) | Yes (NPS) | 3.7 (V) | 0.43 | ********* |
| Suess et al. (2010) (94) | Germany | 51 (129) | 12 (-) | 1 (1) | Influenza A(H1N1)pdm09 | Confirmed | Yes (oseltamivir) | No (no) | Yes (NPA, NPS, OPS) | 3.8 (V) | 1.34 | ****** |
| Thai et al. (2014) (107) | Vietnam | 33 (123) | 16 (-) | 5 (28) | Influenza A(H1N1)pdm09 | Confirmed | Yes (oseltamivir) | Yes (yes) | Yes (NPS) | 3.8 (V) | 1.28 | ******* |
| To et al. (2010) (113) | China | 22 (22) | - | 0 | Influenza A(H1N1)pdm09 | Confirmed | No (pretreatment) | No (no) | Yes (NPA, NPS, OPS) | 3.4 (V) | 0.23 | ***** |
| To et al. (2010) (117) | China | 50 (50) | 0 | 0 | Influenza A(H1N1)pdm09 | Confirmed | Yes (oseltamivir, nebulized zanamivir) | No (no) | Yes (NPA, NPS) | 3.6 (V) | 0.52 | ****** |
| Watanabe et al. (2011) (119) | Japan | 251 (251) | 251 (251) | 0 | Influenza A(H1N1)pdm09 | Confirmed | No (pretreatment) | No (yes) | Yes (NPA) | 3.8 (V) | 2.61 | ********** |
| Wu et al. (2012) (100) | China | 64 (89) | - | 0 | Influenza A(H1N1)pdm09 | Confirmed | Yes (oseltamivir) | No (yes) | Yes (NPS) | 3.7 (V) | 5.53 | ****** |
| Yang et al. (2011) (101) | China | 251 (251) | - | 0 | Influenza A(H1N1)pdm09 | Confirmed | N/A | No (yes) | Yes (OPS) | 3.8 (V) | 6.57 | ***** |

*Data shown as "-" were not obtained from the paper or authors. References before 58 are listed in the main body.
†Responses of "no" for pharmacotherapy are based on no pharmacotherapy given to the patients as described explicitly or none reported in the study.
‡For studies reporting specimen measurements as individual sample data (either in numerical or graphical formats), the sample data was extracted for analysis.
§Specimen measurements were converted to rVLs based on the dilution factor for specimens immersed in transport media.
||Abbreviations for random-effects meta-analyses: virus meta-analysis (V), adult subgroup (A), pediatric subgroup (P), symptomatic/presymptomatic subgroup (S/Ps), asymptomatic subgroup (As).
¶The hybrid JBI Critical Appraisal Checklist was used, with more stars indicating lower risk of bias. Studies were considered to have low risk of bias if they met the majority of the items (≥6/10 items). Results from each study are shown in *SI Appendix*, Table S9.
#For these studies, 2,147 (Lennon et al.) and 134 (Chen et al.) individual specimen measurements were obtained for the individual sample datasets.
☆For Chu et al., only specimen measurements at 20 DFSO were collected, as 5-15 DFSO were specimens reported in Peiris et al.

**Table S7.** Descriptive parameters for respiratory viral loads based on individual sample data.

| | | | Weibull distribution parameters | | Respiratory viral load, $log_{10}$ copies/ml | | | | |
|---|---|---|---|---|---|---|---|---|---|
| **Category** | ***n*** * **(specimens)** | ***n*** * **(studies)** | **Scale factor (95% CI)** | **Shape factor (95% CI)** | **Mean (95% CI)**[†] | **SD**[†] | **80th percentile (95% CI)**[‡] | **90th percentile (95% CI)**[‡] | **99th percentile (95% CI)**[‡] |
| SARS-CoV-2 (overall)[§] | 3.834 | 26 | 7.01 (6.94-7.08) | 3.47 (3.39-3.56) | 6.29 (6.22-6.35) | 2.04 | 8.04 (7.96-8.11) | 8.91 (8.83-9.00) | 10.88 (10.75-11.01) |
| SARS-CoV-1 (overall)[§] | 303 | 5 | 6.37 (6.15-6.60) | 3.40 (3.12-3.71) | 5.72 (5.51-5.93) | 1.86 | 7.33 (7.09-7.57) | 8.14 (7.86-8.43) | 9.98 (9.56-10.42) |
| A(H1N1)pdm09 (overall)[§] | 512 | 10 | 7.39 (7.27-7.51) | 5.43 (5.07-5.81) | 6.81 (6.69-6.94) | 1.45 | 8.07 (7.94-8.20) | 8.62 (8.47-8.76) | 9.79 (9.59-10.00) |
| SARS-CoV-2 (adult)[§] | 3.575 | 20 | 7.00 (6.93-7.07) | 3.48 (3.39-3.57) | 6.27 (6.21-6.34) | 2.03 | 8.02 (7.95-8.10) | 8.89 (8.81-8.98) | 10.86 (10.72-10.99) |
| SARS-CoV-2 (pediatric)[§] | 198 | 9 | 7.43 (7.14-7.74) | 3.63 (3.25-4.05) | 6.69 (6.40-6.97) | 2.06 | 8.47 (8.15-8.80) | 9.35 (8.98-9.73) | 11.32 (10.76-11.90) |
| SARS-CoV-2 (symptomatic/presymptomatic)[§] | 1,574 | 22 | 7.40 (7.30-7.51) | 3.81 (3.67-3.97) | 6.68 (6.58-6.79) | 2.00 | 8.39 (8.28-8.50) | 9.21 (9.09-9.34) | 11.05 (10.86-11.24) |
| SARS-CoV-2 (asymptomatic)[§] | 2,221 | 7 | 6.72 (6.63-6.81) | 3.33 (3.22-3.44) | 6.01 (5.92-6.09) | 2.01 | 8.04 (7.96-8.11) | 8.91 (8.83-9.00) | 10.88 (10.75-11.01) |
| SARS-CoV-2 (all DFSO)[§] | 955 | 21 | 7.07 (6.94-7.21) | 3.50 (3.33-3.68) | 6.35 (6.22-6.48) | 2.03 | 8.10 (7.95-8.25) | 8.97 (8.80-9.15) | 10.94 (10.68-11.21) |
| SARS-CoV-2 (-3 DFSO)[‖] | 1 | 1 | - | - | 10.34 | - | - | - | - |
| SARS-CoV-2 (-2 DFSO)[‖] | 3 | 2 | - | - | 4.22 (2.41-6.02) | 1.59 | - | - | - |
| SARS-CoV-2 (-1 DFSO) | 15 | 5 | 6.17 (5.11-7.47) | 2.82 (1.89-4.19) | 5.48 (4.25-6.70) | 2.21 | 7.31 (6.11-8.75) | 8.30 (6.88-10.02) | 10.62 (8.38-13.45) |
| SARS-CoV-2 (0 DFSO) | 50 | 11 | 6.66 (6.13-7.24) | 3.52 (2.87-4.32) | 6.00 (5.49-6.51) | 1.83 | 7.62 (7.05-8.25) | 8.44 (7.78-9.16) | 10.28 (9.30-11.36) |
| SARS-CoV-2 (1 DFSO) | 63 | 11 | 7.86 (7.33-8.43) | 3.71 (3.04-4.53) | 7.08 (6.54-7.63) | 2.22 | 8.94 (8.36-9.55) | 9.84 (9.17-10.56) | 11.86 (10.84-12.99) |
| SARS-CoV-2 (2 DFSO)[¶] | 71 | 15 | 7.33 (6.84-7.87) | 3.46 (2.85-4.19) | 6.58 (6.07-7.10) | 2.22 | 8.42 (7.87-9.01) | 9.34 (8.69-10.03) | 11.41 (10.39-12.53) |
| SARS-CoV-2 (3 DFSO)[¶] | 75 | 17 | 7.24 (6.73-7.78) | 3.25 (2.70-3.92) | 6.47 (5.95-6.98) | 2.28 | 8.38 (7.81-8.98) | 9.35 (8.68-10.07) | 11.57 (10.52-12.72) |
| SARS-CoV-2 (4 DFSO)[¶] | 85 | 17 | 6.83 (6.29-7.41) | 2.75 (2.32-3.27) | 6.06 (5.54-6.58) | 2.44 | 8.12 (7.51-8.77) | 9.25 (8.52-10.03) | 11.90 (10.72-13.20) |
| SARS-CoV-2 (5 DFSO)[¶] | 93 | 16 | 7.16 (6.69-7.66) | 3.17 (2.69-3.73) | 6.41 (5.95-6.87) | 2.26 | 8.32 (7.80-8.87) | 9.31 (8.70-9.97) | 11.59 (10.63-12.64) |
| SARS-CoV-2 (6 DFSO)[¶] | 105 | 15 | 6.84 (6.41-7.29) | 3.13 (2.67-3.66) | 6.10 (5.68-6.53) | 2.23 | 7.96 (7.49-8.46) | 8.93 (8.36-9.53) | 11.14 (10.24-12.12) |
| SARS-CoV-2 (7 DFSO)[¶] | 136 | 20 | 6.59 (6.23-6.97) | 3.11 (2.71-3.56) | 5.90 (5.55-6.26) | 2.12 | 7.68 (7.27-8.11) | 8.62 (8.14-9.13) | 10.77 (10.05-11.60) |
| SARS-CoV-2 (8 DFSO)[¶] | 123 | 19 | 6.51 (6.12-6.92) | 3.03 (2.62-3.49) | 5.82 (5.44-6.19) | 2.13 | 7.62 (7.18-8.08) | 8.58 (8.06-9.12) | 10.78 (9.96-11.67) |
| SARS-CoV-2 (9 DFSO)[¶] | 128 | 19 | 6.26 (5.87-6.67) | 2.87 (2.50-3.29) | 5.57 (5.20-5.94) | 2.14 | 7.38 (6.95-7.85) | 8.37 (7.85-8.92) | 10.66 (9.83-11.55) |
| SARS-CoV-2 (10 DFSO)[¶] | 115 | 18 | 5.71 (5.30-6.16) | 2.55 (2.20-2.95) | 5.09 (4.70-5.48) | 2.14 | 6.89 (6.41-7.40) | 7.93 (7.35-8.56) | 10.41 (9.45-11.47) |

*These two columns summarize the cumulative number of specimens (left) collected from the number of contributing studies (right) for each category in the systematic dataset.
†The mean and sample SD were calculated on the entirety of individual sample data for each category. These data were collected from studies clearly reporting data for individual specimens.
‡The Weibull quantile distributions were used to determine rVLs at the 80th, 90th and 99th cps.
§These categories included only rVL data from positive (above the detection limit) assay measurements.
‖Data for earlier DFSO were excluded from distribution fitting based on limited data, and empty cells were marked with "-".
¶These categories included negative assay measurements (set at the detection limit to estimate rVLs; *N* = 5, 3, 7, 10, 13, 17, 14, 22 and 17 specimens for 2-10 DFSO, respectively) for cases that tested positive at an earlier DFSO.

**Table S8.** Model parameters describing SARS-CoV-2 kinetics during respiratory infection.

| Parameter | Description | Value (95% CI) | Units |
|---|---|---|---|
| $\beta$ | Infection rate constant | 3.26 (2.21-4.31) | $\times 10^{-7}$ (copies/ml)$^{-1}$ day$^{-1}$ |
| $p$ | Cellular shedding rate of virus | 1.33 (0.74-1.93) | copies/ml day$^{-1}$ cell$^{-1}$ |
| $c$ | Clearance rate of virus | 3.30 (0.25-6.34) | day$^{-1}$ |
| $\delta$ | Clearance rate of infected epithelial cells | 0.71 (0.26-1.15) | day$^{-1}$ |
| $R_{0,c}$ | Cellular basic reproductive number | 9.25 | unitless |

**Table S9.** Assessment of risk of bias based on the hybrid JBI critical appraisal checklist.

| | Checklist items* | | | | | | | | | |
|---|---|---|---|---|---|---|---|---|---|---|
| **Study** | **1** | **2** | **3** | **4** | **5** | **6** | **7** | **8** | **9** | **10** |
| Argyropoulos et al. (76) | | | | | | | | | | |
| Baggio et al. (74) | | | | | | | | | | |
| Fajnzylber et al. (65) | | | | | | | | | | |
| Han et al. (84) | | | | | | | | | | |
| Han et al. (82) | | | | | | | | | | |
| Hung et al. (70) | | | | | | | | | | |
| Hurst et al. (81) | | | | | | | | | | |
| Iwasaki et al. (71) | | | | | | | | | | |
| Kawasuji et al. (85) | | | | | | | | | | |
| L'Huillier et al. (53) | | | | | | | | | | |
| Lavezzo et al. (42) | | | | | | | | | | |
| Lennon et al. (67) | | | | | | | | | | |
| Lucas et al. (75) | | | | | | | | | | |
| Mitja et al. (77) | | | | | | | | | | |
| Pan et al. (83) | | | | | | | | | | |
| Peng et al. (63) | | | | | | | | | | |
| Perera et al. (72) | | | | | | | | | | |
| Shi et al. (69) | | | | | | | | | | |
| Shrestha et al. (68) | | | | | | | | | | |
| To et al. (43) | | | | | | | | | | |
| van Kampen et al. (38) | | | | | | | | | | |
| Vetter et al. (78) | | | | | | | | | | |
| Wölfel et al. (21) | | | | | | | | | | |
| Wyllie et al. (73) | | | | | | | | | | |
| Xu et al. (80) | | | | | | | | | | |
| Yonker et al. (79) | | | | | | | | | | |
| Zhang et al. (62) | | | | | | | | | | |
| Zheng et al. (66) | | | | | | | | | | |
| Zou et al. (64) | | | | | | | | | | |
| Chen et al. (87) | | | | | | | | | | |
| Chu et al. (88) | | | | | | | | | | |
| Chu et al. (89) | | | | | | | | | | |
| Cheng et al. (91) | | | | | | | | | | |
| Hung et al. (90) | | | | | | | | | | |
| Peiris et al. (92) | | | | | | | | | | |
| Poon et al. (86) | | | | | | | | | | |
| Poon et al. (44) | | | | | | | | | | |
| Alves et al. (114) | | | | | | | | | | |
| Chan et al. (105) | | | | | | | | | | |
| Cheng et al. (115) | | | | | | | | | | |
| Cowling et al. (112) | | | | | | | | | | |
| Duchamp et al. (118) | | | | | | | | | | |
| Esposito et al. (109) | | | | | | | | | | |
| Hung et al. (106) | | | | | | | | | | |
| Ip et al. (97) | | | | | | | | | | |
| Ito et al. (108) | | | | | | | | | | |
| Killingley et al. (103) | | | | | | | | | | |
| Launes et al. (102) | | | | | | | | | | |
| Lee et al. (104) | | | | | | | | | | |
| Lee et al. (110) | | | | | | | | | | |
| Li et al. (95) | | | | | | | | | | |
| Li et al. (111) | | | | | | | | | | |
| Loeb et al. (93) | | | | | | | | | | |
| Lu et al. (96) | | | | | | | | | | |
| Meschi et al. (99) | | | | | | | | | | |
| Ngaosuwankul et al. (116) | | | | | | | | | | |
| Rath et al. (98) | | | | | | | | | | |
| Suess et al. (94) | | | | | | | | | | |
| Thai et al. (107) | | | | | | | | | | |
| To et al. (117) | | | | | | | | | | |
| To et al. (113) | | | | | | | | | | |
| Watanabe et al. (119) | | | | | | | | | | |
| Wu et al. (100) | | | | | | | | | | |
| Yang et al. (101) | | | | | | | | | | |

*Descriptions of each item are included in the hybrid JBI critical appraisal checklist (*SI Appendix*, Table S10). References before 58 are listed in the main body. Green, yellow and red represent yes, unclear and no, respectively.

**Table S10.** Hybrid JBI critical appraisal checklist

Reviewer____________________________________Date___________________________

Author______________________________________Year________Record Number_____

| | Yes | No | Unclear | Not applicable |
|---|---|---|---|---|
| 1. Was the sample frame appropriate to address the target population? | ☐ | ☐ | ☐ | ☐ |
| 2. Were the study subjects and the setting described in detail? | ☐ | ☐ | ☐ | ☐ |
| 3. Did the study have consecutive inclusion of participants for case series and cohort studies? Did the study use probability-based sampling for cross-sectional studies? | ☐ | ☐ | ☐ | ☐ |
| 4. Was the response rate adequate, and if not, was the low response rate managed appropriately? | ☐ | ☐ | ☐ | ☐ |
| 5. Was the sample size adequate? | ☐ | ☐ | ☐ | ☐ |
| 6. Were valid methods used for the identification of the condition? | ☐ | ☐ | ☐ | ☐ |
| 7. Were standard, valid methods used for measurement of the exposure? | ☐ | ☐ | ☐ | ☐ |
| 8. Was the exposure measured in an objective, reliable way for all participants | ☐ | ☐ | ☐ | ☐ |
| 9. Was there clear reporting of clinical information of the participants? | ☐ | ☐ | ☐ | ☐ |
| 10. Was statistical analysis appropriate? | ☐ | ☐ | ☐ | ☐ |

Overall appraisal: Include ☐ Exclude ☐ Seek further info ☐

Comments (Including reason for exclusion)

___________________________________________________________________________

___________________________________________________________________________

___________________________________________________________________________

**Tool Guidance**

This hybrid checklist was based on the JBI Critical Appraisal Checklists for case series, prevalence studies and analytical cross-sectional studies.

1. **Was the sample frame appropriate to address the target population?**

   This question relies upon knowledge of the broader characteristics of the population of interest and the geographical area.

   This study broadly investigates the respiratory viral load for the diseases of interest. The population of interest is the general population infected with SARS-CoV-2, SARS-CoV-1, or A(H1N1)pdm09. The geographical area is not constrained. Sample frames restricted to particular subgroups within the general infected population were considered appropriate if they targeted one of the following groups analysed in our study: asymptomatic, presymptomatic, symptomatic, adult, pediatric, hospitalized, non-hospitalized, or community.

2. **Were the study subjects and the setting described in detail?**

   Certain diseases or conditions vary in prevalence across different geographic regions and populations (e.g. Women vs. Men, sociodemographic variables between countries). The study sample should be described in sufficient detail so that other researchers can determine if it is comparable to the population of interest to them

3. **Did the study have consecutive inclusion of participants for case series and cohort studies? Did the study use probability sampling for cross-sectional studies?**

   Inclusion of consecutive participants for case series and cohort studies yields results at lower risk of bias compared to other sampling methods for these study designs. Use of probability-based sampling methods for cross-sectional studies yields estimates at lower risk of bias compared to other sampling methods for this design. Studies that indicate a consecutive inclusion are more reliable than those that do not. For example, a case series that states 'we included all patients (24) with osteosarcoma who presented to our clinic between March 2005 and June 2006' is more reliable than a study that simply states 'we report a case series of 24 people with osteosarcoma.'

4. **Was the response rate adequate, and if not, was the low response rate managed appropriately?**

   A large number of dropouts, refusals or "not founds" amongst selected subjects may diminish a study's validity, as can a low response rates for survey studies. The authors should clearly discuss the response rate and any reasons for non-response and compare persons in the study to those not in the study, particularly with regards to their socio-demographic characteristics. If reasons for non-response appear to be unrelated to the outcome measured and the characteristics of non-responders are comparable to those who do respond in the study, the researchers may be able to justify a more modest response rate.

**5. Was the sample size adequate?**

The larger the sample, the narrower will be the confidence interval around the prevalence estimate, making the results more precise. An adequate sample size is important to ensure good precision of the final estimate. The sample size threshold was calculated as follows:

$$n^* = {z^2\sigma}/{d^2}$$

where $n^*$ is the sample size threshold, $z$ is the z-score for the level of confidence (95%), $\sigma$ is the standard deviation (assumed to be 3 $\log_{10}$ copies/ml, a fourth of the full range of rVLs) and $d$ is the marginal error (assumed to be 1 $\log_{10}$ copies/ml, based on the minimum detection limit for qRT-PCR across studies). This item was met if ≥75% of the included DFSO had ≥46 specimen measurements.

**6. Were valid methods used for the identification of the condition?**

Many health problems are not easily diagnosed or defined and some measures may not be capable of including or excluding appropriate levels or stages of the health problem. If the outcomes were assessed based on existing definitions or diagnostic criteria, then the answer to this question is likely to be yes. If the outcomes were assessed using observer reported, or self-reported scales, the risk of over- or under-reporting is increased, and objectivity is compromised. Importantly, determine if the measurement tools used were validated instruments as this has a significant impact on outcome assessment validity.

**7. Were standard, valid methods used for measurement of the exposure?**

The study should clearly describe the method of measurement of exposure. Assessing validity requires that a 'gold standard' is available to which the measure can be compared. The validity of exposure measurement usually relates to whether a current measure is appropriate or whether a measure of past exposure is needed.

In this study, standard methods to measure viral load in respiratory specimens are RT-PCR quantifying via one of the standard genes for each virus.

**8. Was the exposure measured in an objective, reliable way for all participants?**

The study should clearly describe the procedural aspects of the measurement of exposure as well as factors that can contribute to heterogeneity in measurement.

In this study, objective, reliable interpretation of the exposure depends on the use of quantitative calibration; the specification of extraction; determination of the viral load as a standard metric (e.g., copies/ml or equivalent) or in a manner that can be converted to a standard metric; and, if present, specification of the amount of diluent (e.g., viral transport media) used.

**9. Was there clear reporting of clinical information of the participants?**

There should be clear reporting of clinical information of the participants such as the following information where relevant: disease status, comorbidities, stage of disease, previous interventions/treatment, results of diagnostic tests, etc.

In addition, there should be clear reporting of the number and types (asymptomatic, presymptomatic, symptomatic, adult, pediatric, hospitalized, non-hospitalized, community, etc.) of cases for measurements within the sampling periods of interest. For studies that include data outside of the infectious period, there should be clear reporting of clinical information for participants for the specimen measurements that were collected from within the infectious period.

**10. Was statistical analysis appropriate?**

As with any consideration of statistical analysis, consideration should be given to whether there was a more appropriate alternate statistical method that could have been used. The methods section of studies should be detailed enough for reviewers to identify which analytical techniques were used and whether these were suitable.

| **Risk of bias for each study** | |
|---|---|
| **Low** | The majority of critical appraisal criteria are met (≥6/10 items). The estimates are likely to be correct for the target population. |
| **High** | The majority of critical appraisal criteria are not met (<6/10 items). This may impact on the validity and reliability of the estimates. The estimates may not be correct for the target population. |
| **Unclear** | The majority of items are unclear. There was insufficient information to assess the risk of bias. |
